# Maternal vitamin D status during pregnancy is associated with reduced DNA methylation entropy and hypomethylation of regulatory regions related to immune tolerance

**DOI:** 10.64898/2026.11.16.26368034

**Authors:** Samuel S. Boyd, Julian Hecker, Hooman Mirzakhani, Dawn L. DeMeo, Scott T. Weiss, Arda Halu

## Abstract

**Background:** Vitamin D is crucial to maternal immune system function during pregnancy, acting via the vitamin D receptor (VDR). Yet, there is widespread vitamin D insufficiency among pregnant women. The Vitamin D Antenatal Asthma Reduction Trial investigated maternal vitamin D’s effect on offspring asthma/wheeze. Pregnant women were randomized to a vitamin D supplementation or placebo group, and maternal blood samples were collected at 10-18 and 32-38 weeks of gestation. This analysis aims to identify associations between 25-hydroxyvitamin-D_3_ (25OHD) and DNA methylation (DNAm).

DNAm data were obtained with Illumina’s Infinium Methylation EPICv2 array. Linear mixed effects models assessed DNAm-25OHD associations, adjusting for demographic and clinical factors. Probe-level results were aggregated to identify differentially methylated regions (DMRs), which were annotated to genes via proximity to promoters or enhancers. In addition, DNAm entropy was investigated for associations with 25OHD.

**Results:** Hypomethylated (meso-/hyper-methylated) DMRs showed negative (positive) DNAm-25OHD associations. DMRs overlapped with vitamin D-sensitive topologically associating domains and open chromatin. Annotated genes were primarily enriched for pathways related to antigen processing/presentation. Using paired RNA-seq data, the expression of 59 annotated genes was significantly associated with DNAm of its corresponding DMR. These included *IKBKB*, a kinase with suppressive effects on the NF-kappa-B pathway. Furthermore, DMRs showed negative entropy-25OHD associations, suggesting a more homogeneous immune methylome in women with higher 25OHD levels.

**Conclusions:** Our findings suggest that increased vitamin D may promote methylome stability in genomic regions associated with antigen presentation and immune tolerance, conveying a complex regulatory landscape of the vitamin D-associated methylome during pregnancy.

## Background

A defining characteristic of pregnancy is drastic changes to the maternal immune system, which has the dual mandate of protecting the mother and fetus from pathogens while also tolerating the semi-allogeneic fetus (1, 2). This involves dynamic changes in both innate and adaptive immune functions, including alterations in immune cell composition, cytokine signaling, and inflammatory activity across gestation (1). The proper orchestration of these changes is especially important given fetal plasticity during development and the potential impact of prenatal influences on offspring health (3). Dysregulation of maternal immunological adaptations has been implicated in pregnancy complications such as preeclampsia, preterm birth, gestational diabetes, and offspring asthma (4–6).

In recent decades, vitamin D_3_ (henceforth referred to as vitamin D) has emerged as an important immunomodulator in humans (7). While contained in some foods, the majority of vitamin D is synthesized in the skin via photochemical reactions by ultraviolet radiation. Vitamin D then undergoes 25-hydroxylation to form 25-hydroxyvitamin D_3_ (*i.e.*, 25(OH)D_3_ and henceforth referred to as 25OHD), which is the primary circulating vitamin D metabolite found in humans and the primary metabolite of interest for this study. 25OHD subsequently undergoes 1 -hydroxylation to form 1,25(OH)_2_D_3_, the active form of vitamin D that affects transcriptional regulation by binding with the vitamin D receptor (VDR) transcription factor (TF). Upon binding with 1,25(OH)_2_D_3_, VDR, often as a heterodimer with retinoid X receptor (RXR), interacts with co-activators such as histone acetyltransferases to modify chromatin structure and gene expression (8).

Vitamin D affects both the innate and adaptive immune system (7). Most immune cells express VDR and are able to convert 25OHD into 1,25(OH)_2_D_3_ (9). Antigen presenting cells (APCs), especially dendritic cells (DC), sit at the interface between innate and adaptive immune responses. Studies have shown vitamin D to suppress DC maturation (10, 11) and to promote T regulatory (Treg) cell development (12, 13). In general, these findings point towards a tolerogenic effect of vitamin D on the immune system. One potential mechanism by which vitamin D mediates these effects is the regulation of gene expression via epigenetic modifications, particularly DNA methylation (DNAm) (14, 15).

DNAm, a heritable epigenomic modification with distinct effects on gene transcription, involves the addition of a methyl group to the 5-carbon position of a cytosine nucleotide, catalyzed by DNA methyltransferases. The vast majority of DNAm in humans occurs at cytosine-phosphate-guanine (CpG) dinucleotides (16). CpG-rich regions of the genome form “islands”, often near promoter regions and characterized by hypo-methylation (17). There are studies suggesting that vitamin D is able to promote DNA demethylation, mostly in promoter regions of Vitamin D Receptor (VDR) target genes (14). This demethylation process may occur passively over several cycles of DNA replication, or it may be more active via the recruitment of ten-eleven translocation (TET) enzymes that induce DNA demethylation; however, the underlying mechanisms remain unclear (14).

Our understanding of vitamin D-associated changes to the methylome of pregnant populations is incomplete. We hypothesize that vitamin D may alter DNAm to promote a tolerogenic environment (i.e., one which induces immune tolerance). This epigenome-wide association study (EWAS) aimed to explore associations between maternal DNAm and maternal 25OHD levels from blood samples of pregnant women enrolled in the Vitamin D Antenatal Asthma Reduction Trial (VDAART) (18). We then identified regions of vitamin D-associated methylation and functionally annotated them to genes, regulatory elements, and biological processes. We subsequently assessed the association between DNAm of CpGs and the expression of their annotated genes. Additionally, we investigated the relationship between DNA methylation entropy, an information-theoretic measure that quantifies the heterogeneity in methylation status within a bulk sample, and vitamin D levels (see **Figure 1** for an analysis overview).

**Figure 1.**
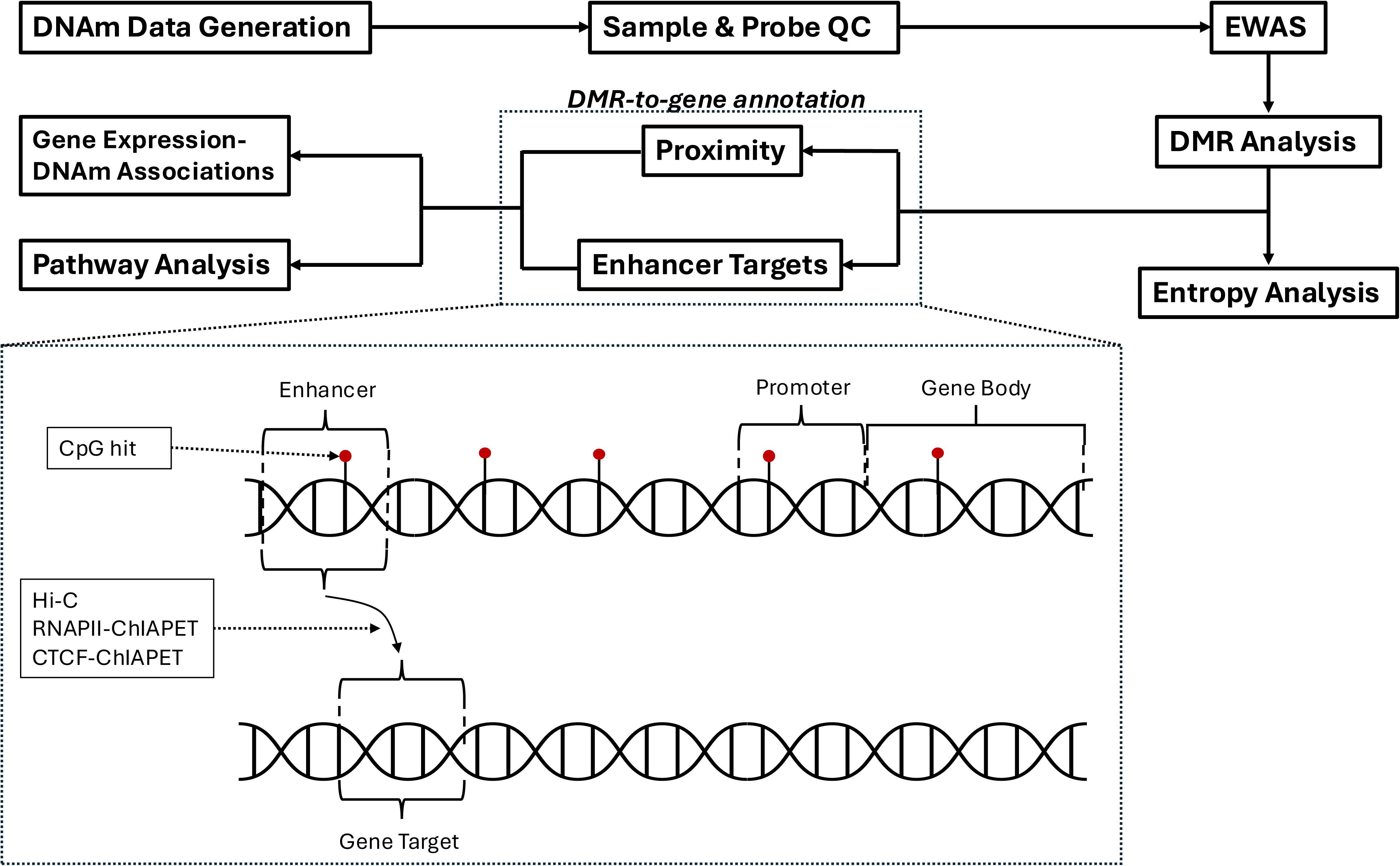
Analysis overview. This flowchart provides the general steps of the analysis for this study. The diagram below illustrates how differentially methylated regions were annotated to genes: via proximity (overlapping a promoter region or gene body), or via enhancers (overlapping an enhancer region which targets one or more genes).

## Results

### Cohort characteristics

**Table 1** shows characteristics of the VDAART intent-to-treat (ITT) cohort, stratified by the existence of DNAm data passing all sample quality control steps. The samples of 477 individuals, of the 806 individuals in the ITT cohort, were assayed for DNA methylation, of which 446 have at least one sample passing all quality control steps (Methods). Sample selection for DNAm assays was restricted to samples with already-extracted DNA. While the majority of cohort characteristics, including demographic variables, treatment group, and maternal BMI, are not significantly different between individuals with and without methylation data, baseline and differential 25OHD levels, age, and collection site differ significantly (**Table 1**). For individuals with methylation data, there are significant differences between the two timepoints for 25OHD levels, vitamin D binding protein (DBP) levels, and season of measurement (**Additional File 1**).

**Table 1.** Baseline characteristics of VDAART intent-to-treat participants with and without DNA methylation data passing all quality control steps. Comparisons between groups was done using a Kruskal-Wallis test for continuous variables, and a Chi-squared test for categorical variables.

|  | DNA methylation data exists |  |  | p-value |
|---|---|---|---|---|
|  | **Yes<br>(N=446)** | **No<br>(N=360)** | **Total<br>(N=806)** |  |
| **Total 25OHD (ng/mL)(M1)** |  |  |  | 0.0006 |
| Mean (SD) | 24.1 (10.8) | 21.5 (9.39) | 23.0 (10.2) |  |
| Median [Min, Max] | 23.3 [5.85, 80.8] | 20.4 [4.35, 56.0] | 22.0 [4.35, 80.8] |  |
| Missing | 1 (0.2%) | 4 (1.1%) | 5 (0.6%) |  |
| **Total 25OHD (ng/mL)(M2)** |  |  |  | 0.6506 |
| Mean (SD) | 33.3 (14.5) | 32.7 (14.8) | 33.0 (14.6) |  |
| Median [Min, Max] | 31.7 [5.67, 90.1] | 31.9 [5.81, 80.2] | 31.7 [5.67, 90.1] |  |
| Missing | 2 (0.4%) | 35 (9.7%) | 37 (4.6%) |  |
| **Total 25OHD (% change, M1 to M2)** |  |  |  | 0.0086 |
| Mean (SD) | 57.1 (91.9) | 69.9 (92.9) | 62.5 (92.5) |  |
| Median [Min, Max] | 35.1 [-77.9, 586] | 46.1 [-62.8, 601] | 39.3 [-77.9, 601] |  |
| Missing | 3 (0.7%) | 38 (10.6%) | 41 (5.1%) |  |
| **DBP (µg/mL)(M1)** |  |  |  | 0.8433 |
| Mean (SD) | 520 (130) | 520 (122) | 520 (129) |  |
| Median [Min, Max] | 517 [178, 1100] | 525 [278, 812] | 518 [178, 1100] |  |
| Missing | 4 (0.9%) | 285 (79.2%) | 289 (35.9%) |  |
| **DBP (µg/mL)(M2)** |  |  |  | 0.4310 |
| Mean (SD) | 674 (204) | 650 (214) | 670 (206) |  |
| Median [Min, Max] | 629 [295, 1750] | 631 [106, 1380] | 630 [106, 1750] |  |
| Missing | 5 (1.1%) | 285 (79.2%) | 290 (36.0%) |  |
| **DBP (% change, M1 to M2)** |  |  |  | 0.2557 |
| Mean (SD) | 33.8 (40.0) | 27.4 (38.8) | 32.9 (39.8) |  |
| Median [Min, Max] | 25.5 [-47.1, 245] | 21.4 [-78.6, 168] | 24.5 [-78.6, 245] |  |
| Missing | 6 (1.3%) | 285 (79.2%) | 291 (36.1%) |  |
| **Maternal Age** |  |  |  | 0.0036 |
| Mean (SD) | 27.9 (5.52) | 26.7 (5.43) | 27.3 (5.51) |  |
| Median [Min, Max] | 28.0 [18.0, 39.5] | 26.8 [18.0, 39.1] | 27.5 [18.0, 39.5] |  |
| **Maternal BMI (M1)** |  |  |  | 0.9337 |
| Mean (SD) | 29.0 (7.75) | 28.9 (7.54) | 28.9 (7.65) |  |
| Median [Min, Max] | 27.3 [16.3, 61.9] | 26.9 [17.3, 54.4] | 27.2 [16.3, 61.9] |  |
| Missing | 14 (3.1%) | 29 (8.1%) | 43 (5.3%) |  |
| **Maternal Asthma Status** |  |  |  | 0.9218 |
| No | 269 (60.3%) | 215 (59.7%) | 484 (60.0%) |  |
| Yes | 177 (39.7%) | 145 (40.3%) | 322 (40.0%) |  |
| **Maternal Income** |  |  |  | 0.0973 |
| Less than \$30,000 | 131 (29.4%) | 105 (29.2%) | 236 (29.3%) |  |
| \$30,000 - \$49,999 | 57 (12.8%) | 48 (13.3%) | 105 (13.0%) |  |
| \$50,000 - \$99,999 | 108 (24.2%) | 68 (18.9%) | 176 (21.8%) |  |
| Over \$100,000 | 56 (12.6%) | 37 (10.3%) | 93 (11.5%) |  |
| Unknown | 94 (21.1%) | 102 (28.3%) | 196 (24.3%) |  |
| **Maternal Ethnicity** |  |  |  | 0.4300 |
| Not Hispanic or Latino | 316 (70.9%) | 265 (73.6%) | 581 (72.1%) |  |
| Hispanic or Latino | 130 (29.1%) | 95 (26.4%) | 225 (27.9%) |  |
| **Maternal Race** |  |  |  | 0.0528 |
| White | 195 (43.7%) | 131 (36.4%) | 326 (40.4%) |  |
| American Indian or Alaskan Native | 3 (0.7%) | 7 (1.9%) | 10 (1.2%) |  |
| Asian | 15 (3.4%) | 21 (5.8%) | 36 (4.5%) |  |
| Black or African American | 181 (40.6%) | 166 (46.1%) | 347 (43.1%) |  |
| Native Hawaiian or Other Pacific Islander | 8 (1.8%) | 3 (0.8%) | 11 (1.4%) |  |
| Other | 44 (9.9%) | 32 (8.9%) | 76 (9.4%) |  |
| **Site** |  |  |  | 0.0207 |
| Boston | 122 (27.4%) | 118 (32.8%) | 240 (29.8%) |  |
| San Diego | 170 (38.1%) | 104 (28.9%) | 274 (34.0%) |  |
| St. Louis | 154 (34.5%) | 138 (38.3%) | 292 (36.2%) |  |
| **Treatment** |  |  |  | 0.1734 |
| Vitamin D Supplementation | 232 (52.0%) | 169 (46.9%) | 401 (49.8%) |  |
| Placebo | 214 (48.0%) | 191 (53.1%) | 405 (50.2%) |  |
| **Offspring Sex** |  |  |  | 0.6236 |
| Female | 217 (48.7%) | 168 (46.7%) | 385 (47.8%) |  |
| Male | 229 (51.3%) | 192 (53.3%) | 421 (52.2%) |  |
| **Offspring asthma/recurrent wheeze by 3 years** |  |  |  | 0.5100 |
| No | 330 (74.0%) | 258 (71.7%) | 588 (73.0%) |  |
| Yes | 116 (26.0%) | 102 (28.3%) | 218 (27.0%) |  |

### Epigenome-wide association study of maternal vitamin D levels reveals differentially methylated CpGs and genomic regions associated with 25OHD

To identify the epigenetic marks associated with maternal vitamin D levels during pregnancy, we conducted an epigenome-wide association study (EWAS) on DNA methylation data derived from maternal blood samples taken at two timepoints during pregnancy. Observations from both timepoints were included in linear mixed effect models to simultaneously capture both between- and within-subject (i.e., across time) effects. These probe-level models revealed two significant CpGs (**Figure 2**). Probe cg17097782_TC11, on chromosome 2, overlaps with several regulatory elements: the gene body of odd-skipped related transcription factor 1 (*OSR1*); a CpG island; a distal enhancer targeting the visinin-like 1 (*VSNL1*) and growth differentiation factor 7 (*GDF7*) genes; and a 25OHD-sensitive open chromatin region. Probe cg11176481_TC21, on chromosome 5, also overlaps with a distal enhancer which targets the forkhead box D1 (*FOXD1*) gene. The genes annotated to these differentially methylated probes (DMP) are enriched in pathways related to bone mineralization, SMAD protein signaling, transforming growth factor *β* (TGF-*β*) signaling, and embryonic organogenesis (**Additional File 2**). See **Additional File 3, Sheets 1-4**, for detailed DMP results. We did not observe any significant interaction effects between gestational age and 25OHD, suggesting that the effects of vitamin D on maternal DNAm are independent of gestational age.

**Figure 2.**
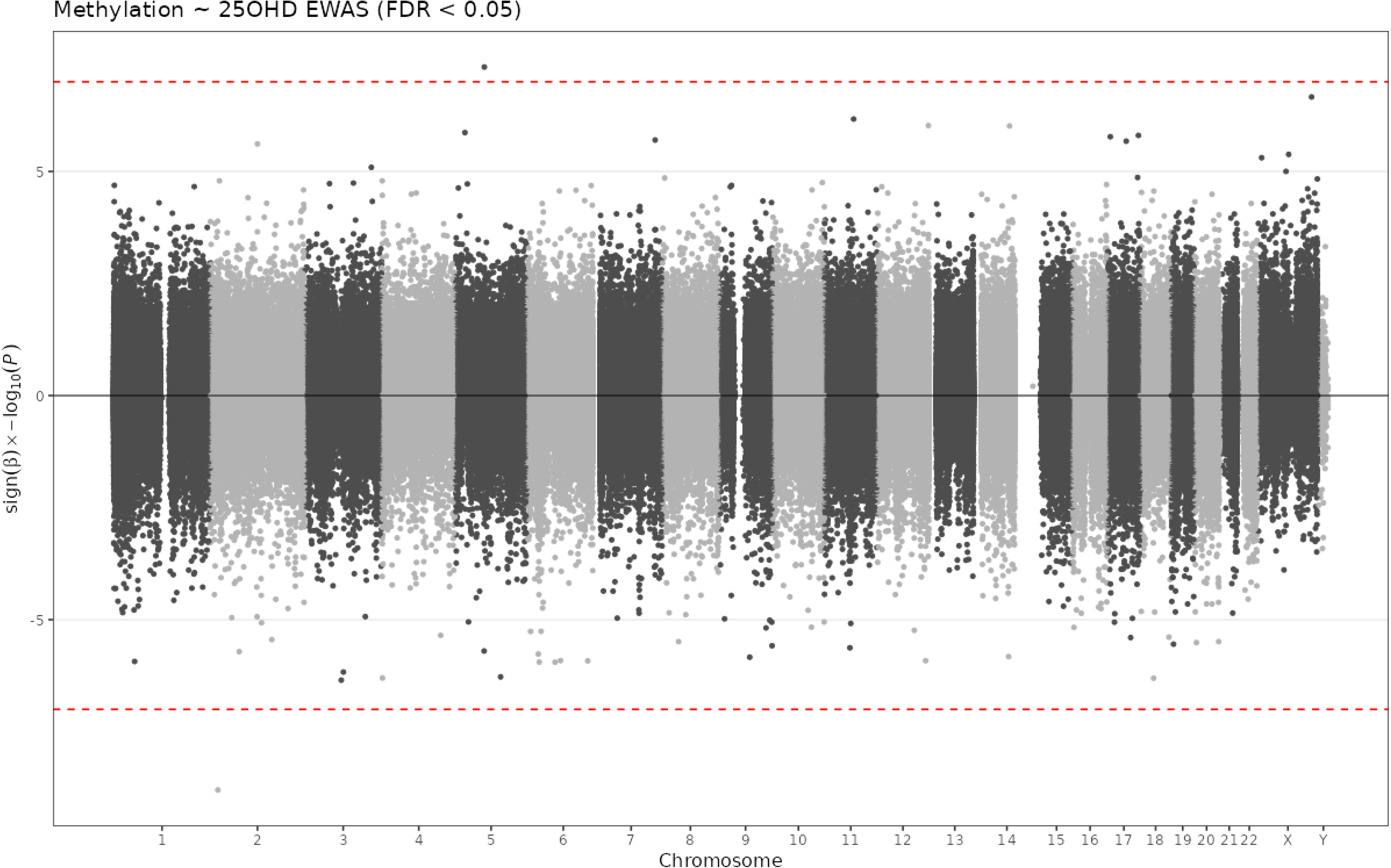
EWAS results. A Manhattan plot of probe-level results from linear mixed models investigating methylation-25OHD associations. The red dashed lines represent a p-value of 4.68*e*-8, corresponding to a false discovery rate of 0.05 for this set of results.

To boost the epigenetic signal and increase statistical power (19), we next conducted a regional DNAm analysis (**Methods**), which resulted in 84 differentially methylated regions (DMRs), containing at least 2 CpGs, with an average region size of 116 bp. A sensitivity analysis shows that the method used to identify DMRs is fairly robust in terms of DMR count, average CpG count, and average region size when varying the distance cutoff and bin size (**Additional File 4**). CpG-level model results for CpGs constituting the identified DMRs can be found in **Additional File 3, sheet 1**. The heatmap in **Figure 3.a** reveals that most hypo-methylated regions are negatively associated with 25OHD, while meso-(intermediate) and hyper-methylated regions show positive associations with 25OHD. **Figure 3.b** further illustrates that methylation levels decreased and increased for hypo- and hyper-methylated DMRs, respectively, from the first to third trimester. These results suggest that an increase in vitamin D levels may stabilize existing methylation regimes in the vitamin D-sensitive parts of the epigenome by lowering DNAm in hypo-methylated DMRs and elevating DNAm in hyper-methylated DMRs.

**Figure 3.**
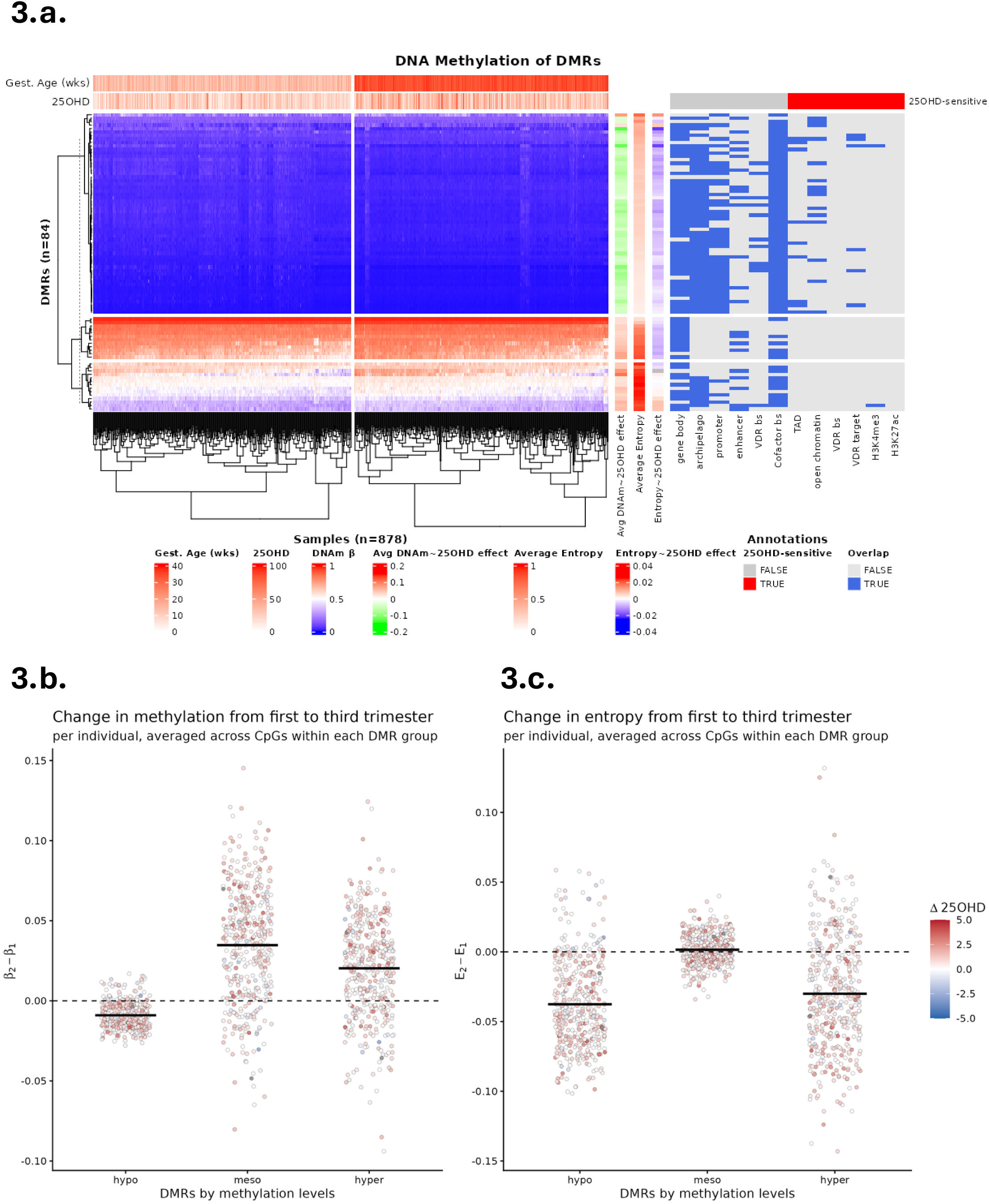
DMR Results. ***a.*** A heatmap of average methylation values per DMR per sample (left), along with various DMR and sample annotations (top, right). At right, a heatmap depicts overlap between DMRs and various classes of regulatory regions. ***b-c.*** Plots of the per-individual change in methylation and entropy levels from the first to third trimester averaged across probes within each DMR group, which are defined by methylation levels (hypo, meso, hyper) from the clustering given in Figure 3.a. Black bars represent the average per group. Dots are colored based on the change in 25OHD levels from the first to third trimester.

### Vitamin D-associated differentially methylated regions map to distinct genomic contexts

Providing genomic context is important to understand the functional implications of the identified epigenetic marks and regions. **Figure 3.a** (right) reveals numerous similarities and contrasts between hypo-, meso-, and hyper-methylated DMRs in terms of overlapping genomic features. All DMRs broadly overlap with gene bodies and co-factor binding sites, and to a large degree, enhancers. On the other hand, hypo-methylated DMRs (in blue) show broad overlap with CpG archipelago and promoter regions, whereas hyper-methylated DMRs (in red) lack any overlap with these regions. Hypo-methylated regions also show a modest overlap with VDR binding sites, in contrast to hyper- and meso-methylated regions. Additionally, meso- and hyper-methylated DMRs show little to no overlap with 25OHD-sensitive regulatory regions, whereas there is some overlap for hypomethylated DMRs, especially with 25OHD-sensitive open chromatin regions. Overall, these patterns may suggest that the downstream regulatory effects of vitamin D are primarily carried out by the hypo-methylated DMRs.

### 25OHD-associated regions are functionally enriched in innate and adaptive immunity, chromatin modeling and cross-presentation-related pathways

The identified DMRs were annotated to 342 unique genes (**Methods**). Of these, 35.4% were mapped via proximity and the rest were mapped via enhancer-target interactions. These genes were significantly enriched in 42 pathways (FDR < 0.05) (**Figure 4**), which clustered into pathway groups that include MHC class I and II antigen processing and presentation, immune response, and ATP-dependent chromatin remodeling. Annotated genes related to antigen presentation include inhibitor of nuclear factor kappa B kinase subunit beta (*IKBKB*) and dynamin 2 (*DNM2*), as well as members of the human leukocyte antigen (HLA), transporter associated with antigen processing (TAP), and proteasome subunit beta (PSMB) gene families. For a complete list of enriched pathways and their associated DMRs and genes, see **Additional File 3, Sheet 7**.

**Figure 4.**
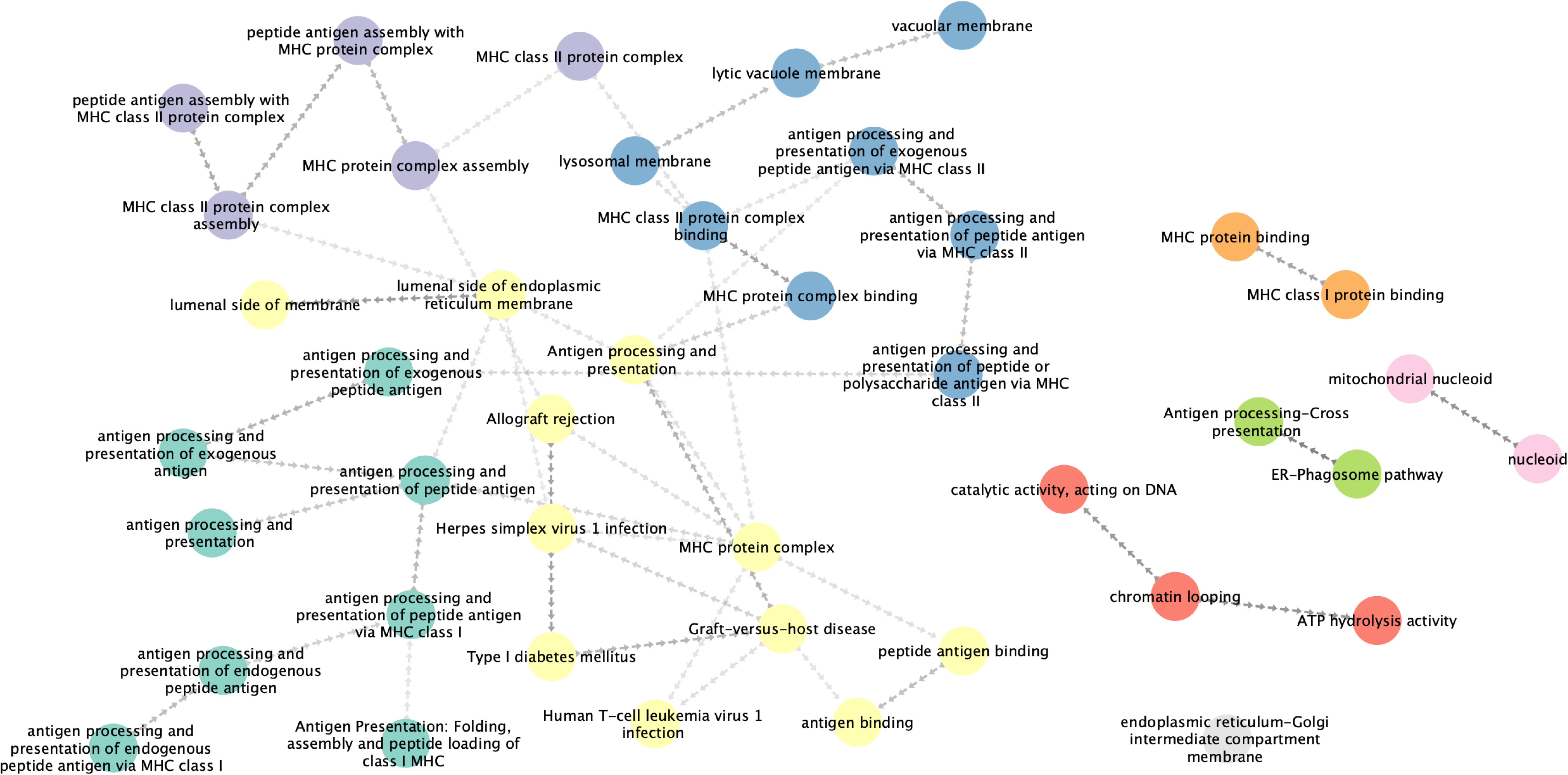
Pathway analysis. Network of pathways significantly enriched for genes annotated to our identified DMRs, using pathways from the Gene Ontology, Reactome, and KEGG databases. Two pathways are connected if the Jaccard Index of their constituent genes is greater than or equal to 0.5, with an arrow pointing from the smaller to the larger pathway. Colors correspond to clusters derived from the Leiden algorithm.

### An increase in vitamin D levels may promote methylation stability in the hypo- and hyper-methylated regions of the maternal epigenome

Our DMR analysis reveals that increased 25OHD levels are associated with decreased methylation in hypo-methylated regions and increased methylation in hyper-methylated regions, which suggests that 25OHD promotes methylation homogeneity (i.e., lower DNAm entropy) at vitamin D-associated regions across cell populations. Motivated by these observations, we hypothesized that the entropy of hypo- and hyper-methylated DMRs (**Methods**) would show negative associations with 25OHD levels.

Most hypo- and some hyper-methylated DMRs show lower average entropy relative to most hyper- and meso-methylated DMRs (**Figure 3.a**), indicating increased methylation homogeneity of cell populations across samples for the former group of DMRs versus heterogeneous methylation patterns for the latter group. As hypothesized, both hypo- and hyper-methylated regions show negative associations between entropy and 25OHD (**Figure 3.a**), which suggests that increased 25OHD levels may reinforce existing methylation regimes, thereby promoting methylation stability (lower entropy) in these regions. We also observe a broad decrease in entropy from first to third trimester for hypo- and hyper-methylated DMRs (**Figure 3.c**). The entropy-25OHD association was significant (FDR < 0.05) for 80 of the 84 DMRs (95%). This association reflects a combination of both between-subject and within-subject (i.e., across time) associations between 25OHD and entropy, which are not distinguished in the model. There were no significant interaction effects between 25OHD and gestational age on entropy. We observed significantly lower entropy for DMRs overlapping archipelago regions, VDR cofactor binding sites, promoters, and 25OHD-sensitive TADs, when compared to DMRs not overlapping these regions (**Figure 5.a**). However, no significant differences were observed for the effect estimates of entropy-25OHD associations when comparing DMRs that do or do not overlap various regulatory regions (**Figure 5.b**). Together, these findings suggest that while entropy of DMRs can help distinguish between different genomic regulatory contexts, this feature does not necessarily generalize to the potential effect of vitamin D on entropy.

**Figure 5.**
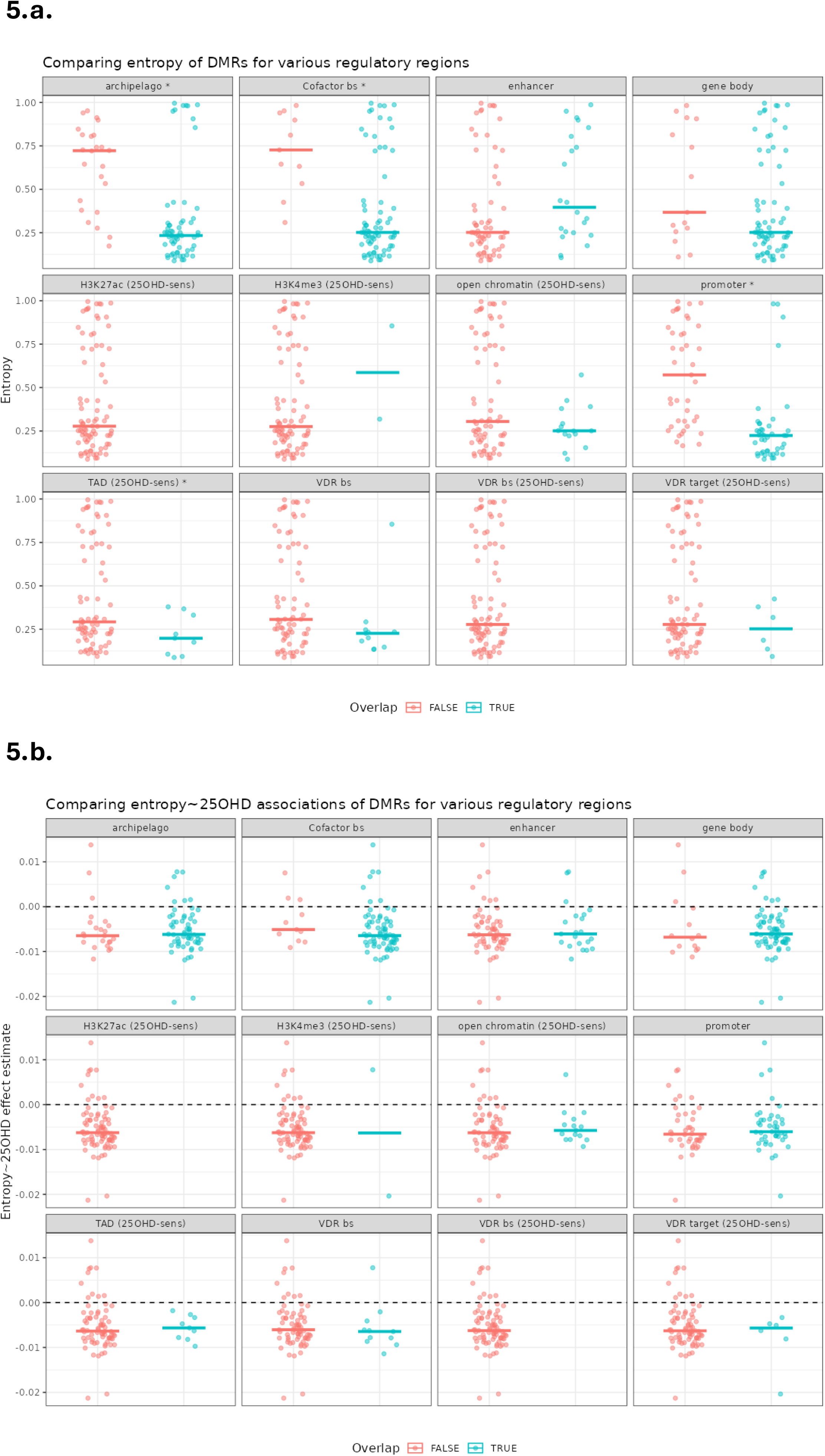
Comparing entropy and entropy-25OHD associations across various regulatory regions. ***a.*** Entropy is compared between overlapping and non-overlapping DMRs for various regulatory regions. For both panels, facets with an asterisk (*) denote significant differences, determined by a Kruskal-Wallis test. ***b.*** The associations between entropy and 25OHD are compared between overlapping and non-overlapping DMRs for various regulatory regions.

To investigate whether our observations on entropy are more extreme than expected by chance, we constructed an empirical background of CpG sets that mimic several properties of our DMRs (**Methods**) and calculated regional entropy (averaged across samples), as well as entropy-25OHD associations, using these background CpG sets. Most of the observed entropy levels for DMRs fall in the low entropy range, although several fall in the high entropy range, whereas the background distribution reveals a bimodal distribution with the primary mode in the mid-entropy range and the secondary mode in the low-entropy range (**Figure 6.a**). For a more global view of the entropy of our identified DMRs, we take the average of the sample-wise averages of entropy over all DMRs and compare this to an empirical background distribution, which shows that it is extremely unlikely to observe by chance a set of 84 regions whose overall average entropy is as low or lower than our observed value (**Figure 6.b**). Finally, when we applied this bootstrap analysis on the estimated effect of 25OHD on regional entropy from linear mixed models, with coefficient estimates weighted by 1 minus the p-value, we found that the majority of the entropy-25OHD effects for the DMRs are negative and fall in the left tail of the empirical distribution (**Figure 6.c**). When considering the average association between entropy and 25OHD across all DMRs, we observe a much stronger negative association than that of randomly chosen sets of 84 regions (**Figure 6.d**).

**Figure 6.**
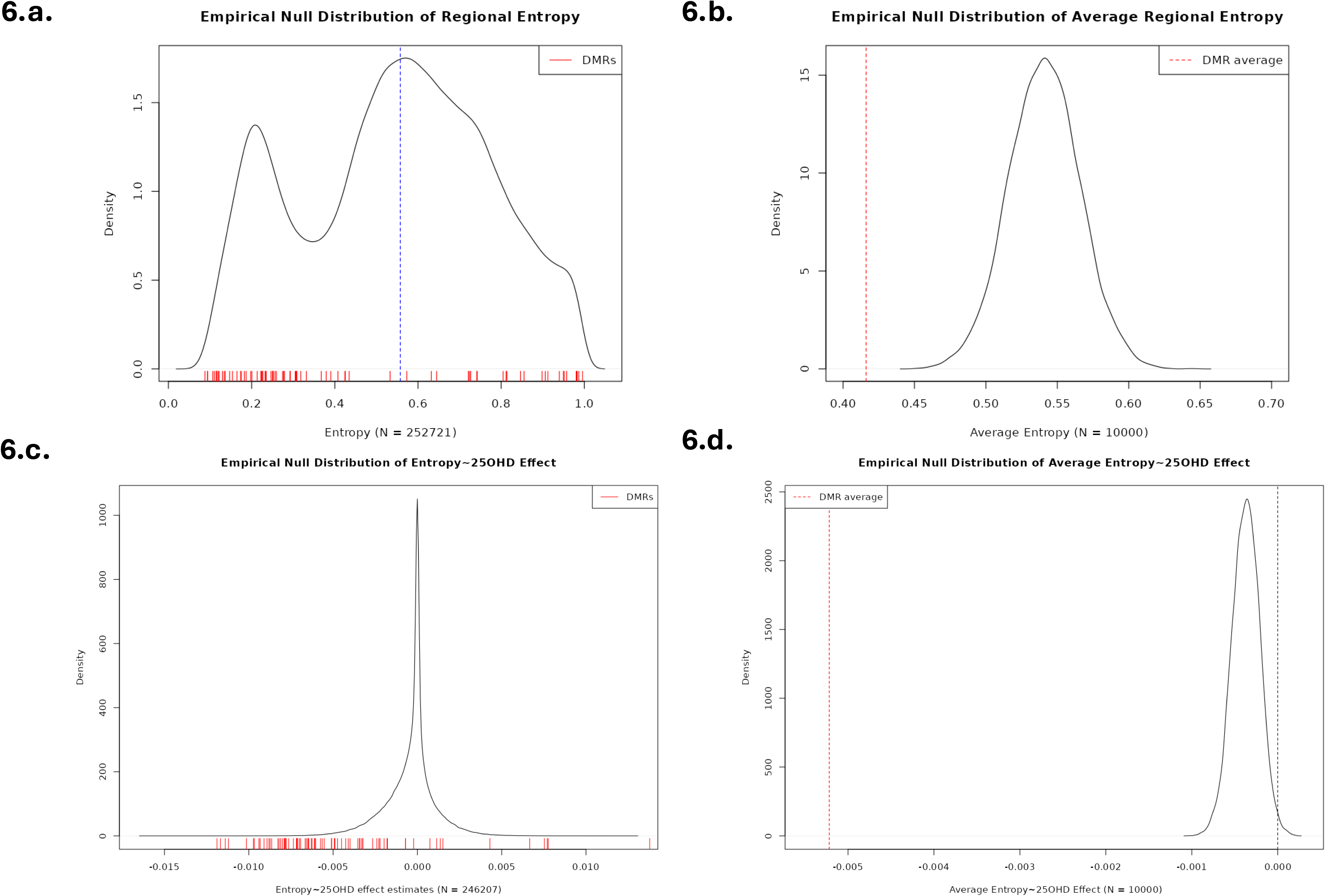
Comparing observed entropy and entropy-25OHD associations of DMRs to null distributions. ***a.*** The entropy of DMRs (red at bottom) is compared to that of random sets of adjacent CpGs (black density curve). Blue dashed line represents the median of null distribution. ***b.*** The average entropy across all DMRs (red dashed line) is compared to the average entropy of randomly drawn sets (n=84) of CpG sets (black density curve). ***c.*** Analogous to panel a. The entropy-25OHD associations (i.e., coefficient estimate weighted by 1 minus p-value) of DMRs (red at bottom) is compared to that of random sets of adjacent CpGs (black density curve). ***d.*** Analogous to panel b. The average entropy∼25OHD association (i.e., coefficient estimate weighted by 1 minus p-value) is compared to that of randomly drawn sets (n=84) of CpG sets (black density curve).

### Integration of gene expression data reveals the potential regulation of maternal-fetal immune tolerance by the maternal vitamin D-associated methylome

The relationship between DNA methylation and gene expression is complex and multi-faceted (20). To investigate the regulatory footprint of the maternal vitamin D-associated methylome, we calculated the associations between the DNAm and gene expression levels of the CpGs in the identified DMRs and their annotated genes, respectively (**Methods**). Of the 501 annotated CpG-gene pairs, 77 had significant associations (p < 0.05) between their DNAm and gene expression levels, with the larger positive effects mainly originating from enhancer-based annotations (**Additional File 5**). The expression-associated DMRs are mostly hypomethylated with negative associations between DNAm and 25OHD levels (**Figure 7.a**). For a given gene, the effect directions are broadly consistent for CpGs within the same DMR, with a nearly even split between positive and negative associations. All but one of the 28 expression-associated DMRs overlap with a CpG island, with the exception overlapping with a CpG shelf. 12 of the 28 expression-associated DMRs overlap with 25OHD-sensitive regions, including TADs, H3K4me3 peaks, VDR target genes, and open chromatin regions.

**Figure 7.**
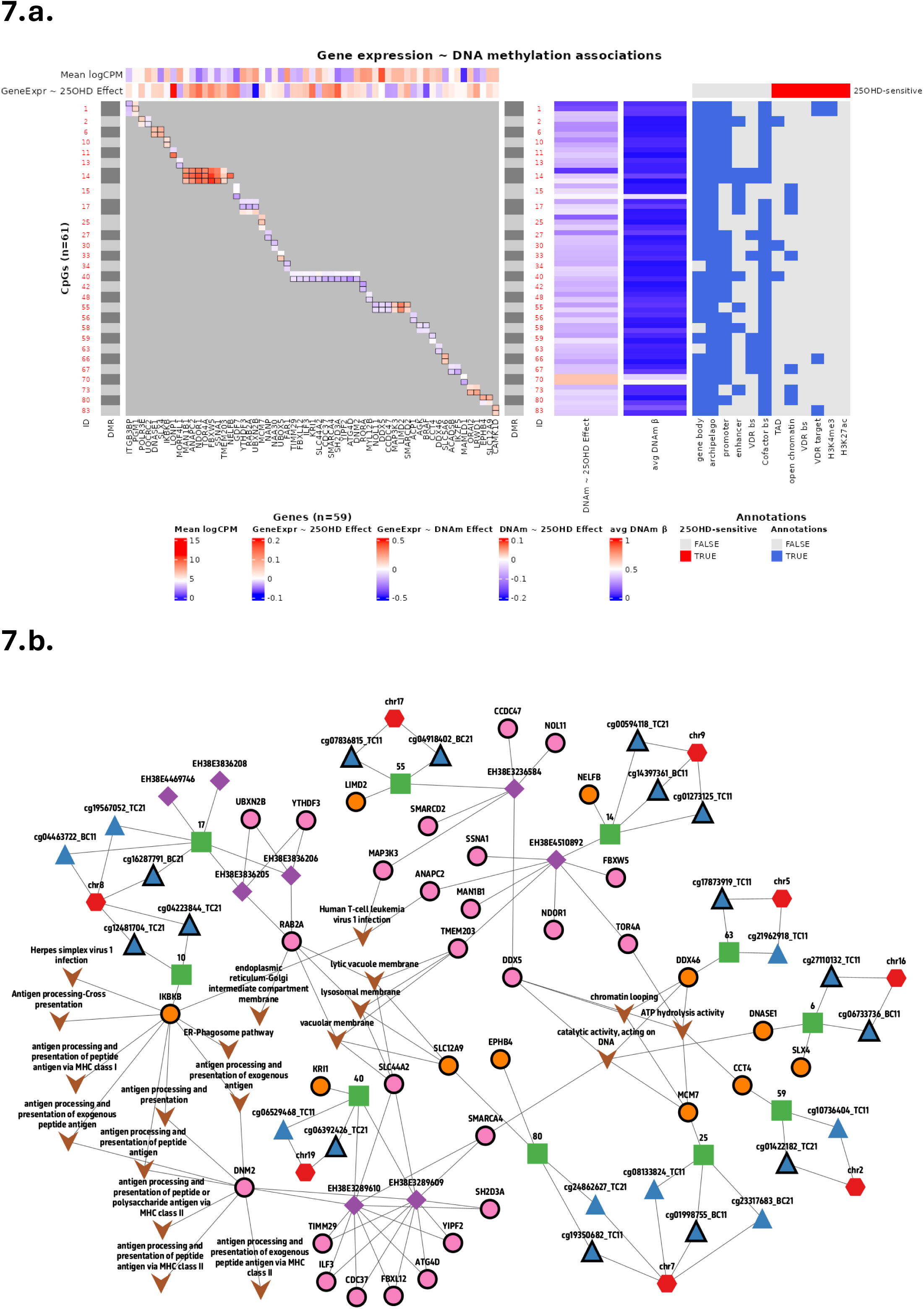
DNA methylation-gene expression associations. ***a.*** A heatmap of CpG-gene associations for all DMRs with at least one constituent CpG showing a significant association with the expression of an annotated gene. Bordered cells represent significant associations (p < 0.05). The heatmap at right depicts DMR overlap with various regulatory elements. ***b.*** Network depicting the relationships between various features used in this study. Red hexagons represent for chromosomes, blue triangles for CpGs, green rectangles for DMRs, purple diamonds for enhancers, orange circles for genes annotated to DMRs via proximity, pink circles for genes annotated to DMRs via enhancers, and brown arrows for significantly enriched pathways. Shown are only DMR-gene pairs with a significant association between DNA methylation and gene expression. CpG-gene pairs with a significant association are represented by both having thick borders with edges connecting them via DMRs (green) and possibly enhancers (purple).

**Figure 7.b** provides an integrated network view of the DMRs, their annotated genes, and the enriched pathways of these annotated genes, focusing on the DMR-gene pairs with significant associations between methylation and gene expression. DMRs 10 and 40 are significantly associated with 1 and 11 genes, respectively, two of which are involved in antigen processing & presentation: *IKBKB* and *DNM2*. *IKBKB* (annotated via proximity; positively associated with methylation of DMR 10) encodes for a kinase which positively regulates the NF-*_k_*B pathway (21, 22). *DNM2* (annotated via enhancer, located in a 25OHD-sensitive TAD; negatively associated with methylation of DMR 40) encodes for a dynamin molecule which facilitates endocytosis of antigen presenting cells (23).

Additionally, **Figure 7.b** reveals that solute carrier family 44 member 2 (*SLC44A2*), solute carrier family 12 member 9 (*SLC12A9*), Ras-related protein rab-2A (*RAB2A*), and transmembrane protein 203 (*TMEM203*), annotated via enhancers to DMRs 40, 80, 17, and 14, respectively, participate in lysosomal/vacuole membrane pathways. On the right side of **Figure 7.b**, several genes are involved in chromatin looping, catalytic activity acting on DNA, and ATP hydrolysis activity, including DEAD-box helicase 5 (*DDX5*; enhancer target of DMR 55; negative association with DNAm) and SWI/SNF related BAF chromatin remodeling complex subunit ATPase 4 (*SMARCA4*; enhancer target of DMR 40; negative association with DNAm). Overall, these results depict a complex regulatory landscape of the vitamin D-associated methylome during pregnancy.

## Discussion

In the present study, we found that 25OHD levels are associated with DNA methylation of sites and regions during pregnancy, which correspond to genes and regulatory regions related to components of maternal-fetal immune tolerance such as TGF-*β* signaling, antigen processing and presentation, and lysosomal membranes. Increased 25OHD levels corresponded to decreased DNAm entropy of hypo- and hyper-methylated regions, while entropy in these regions substantially decreased from the first to third trimester. Furthermore, the DNAm of many of these vitamin D-sensitive regions were found to be significantly associated with the expression of their annotated genes. These findings suggest a functional impact of vitamin D on immune processes, in part mediated by DNAm.

Perhaps the strongest signal from our regional DNAm analysis comes from genomic regions associated with antigen processing and presentation, a critical immune process involving antigen presentation from antigen presenting cells (APCs) along with costimulatory molecules and cytokines to direct T cell differentiation (24). Two DMRs associated with antigen presentation (DMR 10 and 40) show significant associations between their DNAm and the gene expression of their annotated genes: *IKBKB* and *DNM2*. *IKBKB* encodes for a kinase which phosphorylates an inhibitor of the NF-*_k_*B pathway, causing the inhibitor to degrade (21, 22). The NF-*_k_*B pathway is involved in the induction of pro-inflammatory genes in innate immune cells, as well as the development of inflammatory T cells (25, 26). Previous studies have shown that the inhibition of NF-*_k_*B in dendritic cells can lead to down-regulation of MHC-II proteins, co-stimulatory molecules (e.g., CD80, CD40), and pro-inflammatory cytokines (e.g., IL-12, TNF-*α*), thereby promoting immune tolerance (27, 28). DMR 10, whose DNAm was found to have a negative association with 25OHD, lies in exon 1 of *IKBKB*, whose expression was positively associated with DNAm of DMR 10. Taken together, these associations suggest that vitamin D may promote immune tolerance by inhibiting NF-*_k_*B activity via decreased methylation in *IKBKB* and a reduction in *IKBKB* expression.

DMR 40, whose DNAm levels show a negative association with 25OHD levels in our cohort, lies in a distal enhancer that targets *DNM2*, a dynamin molecule involved in antigen presentation which facilitates endocytosis, whose expression is negatively associated with DNAm of the distal enhancer. Our data therefore suggest a positive association between vitamin D and *DNM2* expression. In T cells, *DNM2* has been shown to be critical for actin remodeling of the immunological synapse during antigen presentation, which promotes binding with APCs (29). Therefore, vitamin D may lead to prolonged binding between T cells and APCs, thereby limiting the APC’s ability to detach and activate other T cells (28, 30). Consistent with this hypothesis, a key adhesion molecule, Intercellular adhesion molecule 1 (ICAM1) is also targeted by the distal enhancer overlapping DMR 40 and is negatively associated with DNAm, suggesting a positive relationship between 25OHD and *ICAM1* expression. Moreover, consistent with the vitamin D-mediated inhibition of NF-*_k_*B discussed above, inhibition of NF-*_k_*B in dendritic cells has been shown to up-regulate *ICAM1* (28).

The lysosomal membrane pathway was also significantly enriched for genes identified by our DNAm-25OHD analysis. Initially viewed as an intracellular compartment that facilitates autophagy and recycling of cellular waste, lysosomes have emerged as important signaling platforms, including at the interface between innate and adaptive immunity (31–33). The proteolytic activity of lysosomes is a crucial step for MHC class II antigen presentation and CD4^+^ T cell activation (34). We identified several genes—*TMEM203*, *RAB2A*, *SLC44A2*, and *SLC12A9*, annotated to DMRs 14, 17, 40, and 80, respectively—which participate in the lysosomal pathway in distinct ways (35–38) and whose expression is significantly associated with the DNAm of their corresponding DMR. Our regional DNAm-25OHD analysis therefore implicates two closely related pathways—antigen processing and presentation, and lysosomes—which sit at the intersection of innate and adaptive immunity (39). The exact mechanisms by which vitamin D alters DNAm in these regions and how these alterations might confer immune advantages to mother and fetus during pregnancy remain to be further elucidated.

The TGF-*β* signaling pathway, along with the related SMAD protein signal transduction pathway, was identified as being enriched in annotated genes from our initial probe-level EWAS, primarily the *GDF7* gene, which encodes for a secreted protein that regulates the SMAD family of transcription factors involved in TGF-*β* signaling. TGF-*β* is a cytokine that plays critical roles in regulating the differentiation and function of various immune cell types, including the suppression of Th1/Th2 cell differentiation and the promotion of the generation of peripheral Treg cells (40), which help maintain tolerance towards the allogeneic fetus during pregnancy (6). A DMP (cg17097782_TC11) located on chromosome 2 lies in a distal enhancer targeting the *GDF7* gene, which has been shown to positively regulate Treg cells via increased Forkhead Box P3 (*FOXP3*) expression (41). These findings may suggest that vitamin D promotes a tolerant immune environment via decreased methylation in a distal enhancer targeting the *GDF7* gene, which may result in an increase in *GDF7* expression and the positive regulation of Treg cells.

A major observation from this study is that higher vitamin D associates with decreased methylation in hypo-methylated regions and with increased methylation in hyper-methylated regions. These hypo-methylated DMRs in large part overlap CGIs and promoter regions. Furthermore, a substantial portion of hypo-methylated regions become accessible with vitamin D treatment in monocyte cell lines, which may potentiate treatment-specific epigenetic regulation in these regions. These findings are corroborated by our DNAm entropy analysis, which directly investigated the relationship between vitamin D and the heterogeneity of methylation status across the cell population. For both hypo- and hyper-methylated DMRs, entropy was significantly negatively associated with 25OHD while simultaneously showing large decrease from the first to third trimester. Taken together, these observations may point to a stabilizing effect of vitamin D on methylation patterns in these regions, wherein hypo- and hyper-methylated regimes are reinforced through increased vitamin D levels.

Our additional entropy-based analyses also identified DMRs overlapping CGIs, promoters, VDR cofactor binding sites, and TADs as having significantly lower DNAm entropy compared to non-overlapping DMRs. These results align with previous studies, which found lower DNAm entropy values in CGIs and transcription start sites compared to other genomic regions such as exons, introns, and intergenic regions (42, 43). Our results show decreased DNAm heterogeneity (i.e., lower DNAm entropy) across cell populations for key regulatory elements involved in chromatin structure and gene transcription. Additionally, the DNAm entropy of identified DMRs was found to be lower than expected by chance when compared to random regions of comparable size and CpG density. These results indicate that genomic regions whose DNAm levels associate with 25OHD levels show significantly higher homogeneity in methylation status across cell populations.

The present study comes with several limitations. The Illumina V2 epigenome chip covers 935,000 CpG methylation sites or 3.3% of the human epigenome centered on CpG islands but certainly not the whole epigenome. Our blood samples were assayed in bulk, thus preventing a single-cell resolution and requiring deconvolution of cell type proportions to control for confounding in statistical models. Additionally, there are fewer than 100 samples with paired DNAm and RNA-seq data, which limits statistical power and the generalizability of observed DNAm-gene expression relationships. EWAS as a statistical methodology may be underpowered to detect all of the relevant CpG-25OHD associations. Further studies with whole-genome sequencing, single-cell resolution of DNAm, ideally coupled with data on gene expression and chromatin structure, would allow researchers to distinguish between immune cell types when investigating the effects of vitamin D on methylation dynamics during pregnancy. Our study is of modest sample size and also lacks a replication cohort, which warrants caution regarding the generalizability of our findings to other populations of pregnant women.

## Conclusions

In conclusion, we found that DNAm patterns in pregnant women associate with vitamin D levels, localizing to hypo- and hyper-methylated genomic regions associated with key processes at the intersection of the innate and adaptive immune systems, primarily antigen processing and presentation and lysosomal pathways. The expression of several genes corresponding to these pathways were also correlated with DNAm levels. Furthermore, these vitamin D-associated regions show significantly higher homogeneity in methylation status across cell populations. Our findings suggest that increased vitamin D may promote methylome stability in genomic regions associated with antigen processing and presentation, potentially promoting immune tolerance towards the allogeneic fetus during pregnancy.

## Methods

### VDAART Design

VDAART is a randomized, double blind, placebo-controlled clinical trial of vitamin D supplementation (4,000 IU plus a multivitamin with 400 IU daily) versus placebo (placebo pill plus a multivitamin with 400 IU daily) in pregnant women, given after baseline measurements were taken, to prevent adverse pregnancy outcomes including asthma and recurrent wheeze in their children. Eligible participants were women between the ages of 18 and 39 years with the estimated enrollment gestational ages between 10 and 18 weeks (singleton) who had a history of physician-diagnosed asthma or atopy, or a partner (biologic father of the child) with a history of physician-diagnosed asthma or atopy, and no history of smoking or current nicotine product use. Eligible participants were screened per VDAART protocol and enrolled if eligible. A participant flow chart is available in **Additional File 6**. Details of the trial design, protocol, and primary and secondary outcomes have been published previously (18, 44, 45). For blood sample collection and processing details and covariate assessment details, see (18, 44). VDAART was approved by the Institutional Review Boards of the participating clinical centers and the data coordinating center, with pregnant women signing informed consent at the enrollment visit covering both primary and secondary data analyses. In this study, we included pregnant women enrolled in VDAART who provided any postnatal data, up to the child’s age of 3 years, and who had DNAm data that passed quality control steps (**Table 1**).

### DNA methylation data processing

Blood samples were drawn from pregnant women participating in VDAART at the first (10-18 weeks of gestation) and third (32-38 weeks of gestation) trimesters (referred to as M1 and M2, respectively, throughout the text) and stored at -80°C. DNA was extracted from 947 blood samples from 477 individuals (477 samples from M1, 470 samples from M2, 470 overlap), using the EZ DNA Methylation-Gold Kit (Zymo Research, Irvine, CA). Samples were randomized across slides, while keeping samples from the same individual together on the same slide. The Illumina Infinium Methylation EPIC v2 array (Illumina, San Diego, CA) was used to perform genome-wide methylation assays of bisulfite-converted DNA samples across a total of 936,990 CpG sites. Pre-processing was performed using R programming language v4.5.1 (R Core Team, 2025) and the *minfi* package v1.54.1 (46).

The sample quality control steps included removing technical replicates, samples with failed bisulfite conversion, samples with genotype or sex mismatches, and outlier samples. This resulted in the removal of 62 of 947 samples, for a total of 885 retained samples (**Additional File 7**). The probe quality control steps included removing probes with poor quality or non-unique mapping; probes with large detection p-values; probes labeled as “new variant” (without an rsID); and probes whose body, targeted CpG site, or single base extension overlapped with a common single nucleotide polymorphism (SNP) with minor allele frequency 5%. This resulted in the removal of 124,320 of 936,990 probes, for a total of 812,670 retained probes (**Additional File 7**).

The normal-exponential out-of-band method (*minfi::preprocessNoob* function, following the *package::function* notation used in R) was applied for background correction and dye-bias normalization. The functional normalization method (*minfi::preprocessFunnorm*) was then used for between-array normalization. Finally, methylation values were logit-transformed (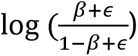 with offset term *ε*) to obtain so-called M-values, which were used as input for linear modeling.

### DNA methylation data analysis

Linear mixed effects models were used to assess the relationships between probe-level DNAm M-values and 25OHD levels (ng/mL), with DNAm as the dependent variable. The models were adjusted for maternal age, BMI at first trimester, gestational age at sample collection, sample collection site, maternal asthma status, vitamin D treatment randomization group, season of sample collection, the first five ancestry principal components derived from VDAART genotype data (see (47) for details), and cell type proportion estimates. Illumina array slide and participant ID were included as random intercepts. Additionally, an interaction term between 25OHD and gestational age was included, to assess if the DNAm-25OHD relationship changes during pregnancy. Cell type proportions were estimated at each timepoint via *EpiDISH::epidish* in R (48), using robust partial correlation and a whole blood reference dataset of DNAm markers of 12 blood cell types for the EPIC array (49). The log ratios of cell type proportion estimates, with neutrophils as the referent, were included as covariates in linear mixed effects models. The function *dream* from the *variancePartition* package in the R programming language was used to fit the linear mixed effects models, using restricted maximum likelihood for model fitting and the Satterthwaite method for the estimation of effective degrees of freedom for hypothesis testing (50).

Linear mixed effects model results of adjacent CpGs were aggregated via the *Enmix::ipdmr* function in R, to identify differentially methylated regions (DMRs) (51). Single probes were excluded. Default function arguments for *Enmix::ipdmr* were used, including a region-level FDR threshold of 0.05, to identify DMRs.

### Differentially methylated region annotations

The CpG sets representing DMRs were annotated to genes in two ways: via proximity and via enhancer targets (**Figure 1**). For the proximity annotation, DMRs were annotated to genes if they overlap with either the promoter region or the gene body. For the enhancer annotation, enhancer-like regulatory elements and their putative gene targets were obtained from SCREEN v4 (52). DMRs overlapping with an enhancer region were annotated to the gene targets of that enhancer.

DMRs were further annotated to additional putative regulatory elements: promoter regions; CpG islands, shores, and shelves (henceforth denoted collectively as the “archipelago”); VDR bindings sites (BS); and VDR co-factor BS. Putative promoter regions were obtained from the SCREEN database and are defined as regions located near transcription start sites (TSS) with high chromatin accessibility and H3K4me3 signal, with data restricted to those originating from blood-based cell types. CpG archipelago annotations were obtained from Illumina’s manifest file on (EPIC v2.0 A2). VDR and VDR co-factor binding sites were obtained from ChIP Atlas, while also restricting data to those originating from blood-based cell types (53). A transcription factor (TF) was considered a potential co-factor of VDR if it has an average co-localization score greater than 1 (more details at https://github.com/inutano/chip-atlas/wiki#colocalization_doc, accessed 2026/06/01).

The binding of VDR with 1,25OHD affects how VDR interacts with the genome and how it modifies chromatin structure. Therefore, data on 25OHD-sensitive regions from THP-1 cell lines (human monocyte) were obtained from publicly available sources to further investigate vitamin D-responsive regulatory elements. Genomic features showing significant differences between 25OHD-stimulated and control samples include H3K27ac and H3K4me3 histone marks (Table S2 in (54)), open chromatin (Supplementary Data File 008 in (55)), topologically associating domains (TADs)(Table S3 in (56)), VDR binding sites (Table S2 in (57)), and VDR target genes (Table S4 in (57)).

### Gene expression data processing and quality control

Raw whole-blood mRNA-seq reads were processed using bcbio-nextgen (version 1.2.9). Read quality was assessed using FastQC (version 0.11.9), followed by adapter trimming and quality filtering with cutadapt (version 3.4). Processed reads were aligned to the GRCh38 human reference genome using STAR (version 2.7.10a). Gene-level counts were generated using featureCounts from the Subread package (version 2.0.3), based on GENCODE version 46 gene annotations. Quality-control metrics included sequencing depth, alignment rate, duplicate rate, and ribosomal RNA content.

Sample-level expression profiles were examined for potential outliers using multidimensional scaling, implemented with the plotMDS function in the limma R package. Three clear outliers, one collected during early pregnancy (M1) and two collected during late pregnancy (M2), were identified and excluded. Samples without corresponding DNA methylation data were also excluded. The final analytic dataset included 152 samples from 89 participants, with 73 samples collected at M1 and 79 at M2.

Lowly expressed features were removed using filterByExpr with default settings in edgeR (version 4.6.3) (58), resulting in a final expression dataset comprising 14,899 expressed genes. TMM normalization factors were estimated using the edgeR calcNormFactors function to account for differences in library size and RNA composition.

### Gene expression data analysis

The filtered RNA-seq count data and TMM normalization factors were supplied to voomWithDreamWeights in the variancePartition R package to estimate the mean–variance relationship and generate precision weights for log2 counts per million. Gene-wise linear mixed-effects models were subsequently fitted using the variancePartition dream function.

For each annotated CpG–gene pair, a gene-wise linear mixed-effects model was fitted using the variancePartition dream function, with gene expression specified as the outcome and DNA methylation as the primary predictor. Models included maternal age, BMI at enrollment, gestational age at sample collection, study site, maternal asthma status, randomized treatment group, the first five genotype principal components, and estimated blood-cell proportions as fixed-effect covariates. Illumina array slide and participant ID were included as random intercepts. The participant-specific random intercept accounted for the correlation between repeated M1 and M2 samples from the same participant.

Associations between circulating 25OHD concentrations and gene expression were evaluated separately using linear mixed-effects models, with gene expression specified as the outcome and 25OHD concentration as the primary predictor. These models included the same covariates and a participant-specific random intercept as specified in the previous paragraph, but excluded Illumina array slide.

### Pathway Analysis

Genes obtained from DMR annotation were assessed for their enrichment in biological pathways from Gene Ontology (GO), Reactome, and KEGG databases (59–61). Using annotated genes as the query set and all known genes (GENCODE v49 on genome build hg38) as the background set, over-representation analysis was performed using a hypergeometric test with an FDR threshold of 0.05. The R packages *clusterProfiler* and *ReactomePA* were used for pathway analysis (62, 63). To construct a network visualization of significant pathways, edges were created between two pathways if the Jaccard Index of their constituent genes is greater than or equal to 0.5, with node colors corresponding to clusters derived from the Leiden algorithm (64).

### Entropy Analysis

Blood samples in this study were processed in bulk for DNAm data generation. Therefore, the obtained DNAm *β* values can be viewed as the fraction of cells from the bulk sample in which a given CpG site is methylated. By treating *β* values as Bernoulli random variables, we compute the entropy for probe *i*, sample *j*, as follows:

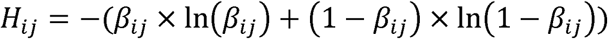

For a set of probes, the standardized average entropy for probe set *P_k_*, sample *j*, is calculated as follows:

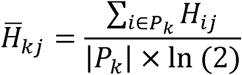

The standardized average entropy (referred to simply as “entropy” throughout the text) measures the degree of heterogeneity in methylation status for a set of probes across the cell population of a bulk sample, and it varies between 0 and 1 (65). Low entropy (i.e., low heterogeneity in methylation status across a cell population) represents DNAm *β* values close to 0 or 1, whereas high entropy represents DNAm *β* values close to 0.5.

Linear mixed effects models were used to assess the relationships between standardized average entropy of CpG sets and 25OHD levels, with entropy as the dependent variable. The modeling is analogous to the DNAm-25OHD models. The models were adjusted for maternal age, BMI at first trimester, gestational age at sample collection, sample collection site, maternal asthma status, randomization group, season of sample collection, the first five principal components of genotype data, and cell type proportion estimates. Illumina array slide and participant ID were included as random intercepts. Additionally, an interaction term between 25OHD and gestational age was included, to assess if the entropy-25OHD relationship changes during pregnancy.

The CpG sets used to calculate entropy and to fit models were first derived from our identified DMRs. To compare the DMR results to those of random regions, the probes on the Infinium EPIC v2 array passing quality control steps were exhaustively searched for sets of adjacent probes spanning at most 1kb and containing either two or three probes (matching parameters and results from *ipDMR* method). These probe sets were then used to calculate an empirical background distribution of regional entropy (averaged across samples) as well as entropy-25OHD associations. Additionally, for a given statistic of interest (e.g., average entropy, entropy-25OHD associations), we take the average over all DMRs and compare this to an empirical background distribution generated by sampling 84 of the 252,721 background CpG sets (without replacement) and calculating the average statistic, repeating this sampling procedure 10,000 times with replacement.

## Declarations

## Ethics approval and consent to participate

VDAART was approved by the Institutional Review Boards of the participating clinical centers and the data coordinating center (MGB IRB protocol number 2009P000557), with pregnant women signing informed consent at the enrollment visit covering both primary and secondary data analyses. This secondary analysis was conducted in accordance with the ethical principles outlined by the NIH and the institutional review board of MGB (IRB protocol number 2023P003089).

## Consent for publication

Not applicable.

## Availability of data and materials

Data described in the manuscript, code book, and analytic code will be made available upon request pending application and approval.

## Authors’ contributions

SSB contributed to the design of the study, performed the statistical analyses, and drafted the manuscript. HM, DLD, and STW contributed to the design of the study and provided critical revisions. HM also contributed to the acquisition of data. AH contributed to the conception and design of the study and the acquisition of data and provided critical revisions. All authors approve of the final manuscript.

## Acknowledgements

Not applicable.

## Additional Files

**Additional File 1.** Comparison, across timepoints, of samples with DNA methylation data passing all quality control steps. Comparisons between groups was done using a Kruskal-Wallis test for continuous variables, and a Chi-squared test for categorical variables.

**Additional File 2.** *DMP-level pathway analysis*. Network of pathways significantly enriched for genes annotated to our identified DMPs, using pathways from the Gene Ontology, Reactome, and KEGG databases. Two pathways are connected if the Jaccard Index of their constituent genes is greater than or equal to 0.5. Colors correspond to clusters derived from the Leiden algorithm.

**Additional File 3.** *Sheet 1*. DNAm-25OHD model results. This includes all constituent CpGs of our identified DMRs, as well as statistically significant CpGs from linear mixed effects models. *Sheet 2*. DMP regulatory annotations. *Sheet 3*. DMP-to-gene annotations. *Sheet 4*. DMP pathway analysis. *Sheet 5*. DMR results. This includes genomic location, constituent CpGs, and regulatory region overlap. *Sheet 6*. DMR-to-gene annotations. *Sheet 7*. DMR pathways analysis. *Sheet 8*. Gene expression-DNAm model results. This includes results for all CpG-gene pairs identified from DMR-to-gene annotations.

**Additional File 4.** *ipDMR sensitivity analysis*. The ipDMR method for identifying differentially methylated regions (DMRs) was assessed—in terms of DMR count (left), average probe count (middle), and average region size (right)—for its sensitivity to its input parameters: CpG distance cutoff; bin size for calculating covariance among CpGs; FDR threshold; and the inclusion of CpG singletons.

**Additional File 5.** *Volcano plot of DNA methylation-gene expression associations*. A volcano plot of linear mixed model results for gene expression ∼ DNA methylation associations. Red dashed line represents a p-value of 0.05.

**Additional File 6.** *Vitamin D Antenatal Asthma Reduction Trial Participant Exclusion Flowchart*. This flowchart shows the reasons for participant exclusion from this study, from the initial eligibility assessment of the Vitamin D Antenatal Asthma Reduction Trial to DNA methylation data generation from maternal blood samples.

**Additional File 7.** *Sample and probe quality control*. ***a.*** Flowchart showing reasons for the exclusion of samples from this study. ***b.*** Flowchart showing reasons for the exclusion of Illumina Infinium EPIC v2 array probes from this study.

## References

1. Abu-Raya B, Michalski C, Sadarangani M, Lavoie PM. Maternal Immunological Adaptation During Normal Pregnancy. Front Immunol. 2020;11:575197.

2. Yang F, Zheng Q, Jin L. Dynamic Function and Composition Changes of Immune Cells During Normal and Pathological Pregnancy at the Maternal-Fetal Interface. Front Immunol. 2019;10:2317.

3. Calkins K, Devaskar SU. Fetal origins of adult disease. Curr Probl Pediatr Adolesc Health Care. 2011;41(6):158–76.

4. DeVries A, McCauley K, Fadrosh D, Fujimura KE, Stern DA, Lynch SV, et al. Maternal prenatal immunity, neonatal trained immunity, and early airway microbiota shape childhood asthma development. Allergy. 2022;77(12):3617–28.

5. Kalagiri RR, Carder T, Choudhury S, Vora N, Ballard AR, Govande V, et al. Inflammation in Complicated Pregnancy and Its Outcome. Am J Perinatol. 2016;33(14):1337–56.

6. Munoz-Suano A, Hamilton AB, Betz AG. Gimme shelter: the immune system during pregnancy. Immunol Rev. 2011;241(1):20–38.

7. Bikle DD. Vitamin D metabolism, mechanism of action, and clinical applications. Chem Biol. 2014;21(3):319–29.

8. Seuter S, Pehkonen P, Heikkinen S, Carlberg C. Dynamics of 1alpha,25-dihydroxyvitamin D3-dependent chromatin accessibility of early vitamin D receptor target genes. Biochim Biophys Acta. 2013;1829(12):1266–75.

9. Hewison M. Vitamin D and immune function: an overview. Proc Nutr Soc. 2012;71(1):50–61.

10. Adorini L, Penna G, Giarratana N, Uskokovic M. Tolerogenic dendritic cells induced by vitamin D receptor ligands enhance regulatory T cells inhibiting allograft rejection and autoimmune diseases. J Cell Biochem. 2003;88(2):227–33.

11. Penna G, Adorini L. 1 Alpha,25-dihydroxyvitamin D3 inhibits differentiation, maturation, activation, and survival of dendritic cells leading to impaired alloreactive T cell activation. J Immunol. 2000;164(5):2405–11.

12. Jeffery LE, Burke F, Mura M, Zheng Y, Qureshi OS, Hewison M, et al. 1,25-Dihydroxyvitamin D3 and IL-2 combine to inhibit T cell production of inflammatory cytokines and promote development of regulatory T cells expressing CTLA-4 and FoxP3. J Immunol. 2009;183(9):5458–67.

13. Urry Z, Xystrakis E, Richards DF, McDonald J, Sattar Z, Cousins DJ, et al. Ligation of TLR9 induced on human IL-10-secreting Tregs by 1alpha,25-dihydroxyvitamin D3 abrogates regulatory function. J Clin Invest. 2009;119(2):387–98.

14. Fetahu IS, Hobaus J, Kallay E. Vitamin D and the epigenome. Front Physiol. 2014;5:164.

15. Ong LTC, Booth DR, Parnell GP. Vitamin D and its Effects on DNA Methylation in Development, Aging, and Disease. Mol Nutr Food Res. 2020;64(23):e2000437.

16. Greenberg MVC, Bourc’his D. The diverse roles of DNA methylation in mammalian development and disease. Nat Rev Mol Cell Biol. 2019;20(10):590–607.

17. Angeloni A, Bogdanovic O. Sequence determinants, function, and evolution of CpG islands. Biochem Soc Trans. 2021;49(3):1109–19.

18. Litonjua AA, Carey VJ, Laranjo N, Harshfield BJ, McElrath TF, O’Connor GT, et al. Effect of Prenatal Supplementation With Vitamin D on Asthma or Recurrent Wheezing in Offspring by Age 3 Years: The VDAART Randomized Clinical Trial. JAMA. 2016;315(4):362–70.

19. Jaffe AE, Murakami P, Lee H, Leek JT, Fallin MD, Feinberg AP, et al. Bump hunting to identify differentially methylated regions in epigenetic epidemiology studies. Int J Epidemiol. 2012;41(1):200–9.

20. Wagner JR, Busche S, Ge B, Kwan T, Pastinen T, Blanchette M. The relationship between DNA methylation, genetic and expression inter-individual variation in untransformed human fibroblasts. Genome Biol. 2014;15(2):R37.

21. Ghosh S, Karin M. Missing pieces in the NF-kappaB puzzle. Cell. 2002;109 Suppl:S81–96.

22. Liu T, Zhang L, Joo D, Sun SC. NF-kappaB signaling in inflammation. Signal Transduct Target Ther. 2017;2:17023-.

23. Gold ES, Underhill DM, Morrissette NS, Guo J, McNiven MA, Aderem A. Dynamin 2 is required for phagocytosis in macrophages. J Exp Med. 1999;190(12):1849–56.

24. Roche PA, Furuta K. The ins and outs of MHC class II-mediated antigen processing and presentation. Nat Rev Immunol. 2015;15(4):203–16.

25. Lawrence T. The nuclear factor NF-kappaB pathway in inflammation. Cold Spring Harb Perspect Biol. 2009;1(6):a001651.

26. Tak PP, Firestein GS. NF-kappaB: a key role in inflammatory diseases. J Clin Invest. 2001;107(1):7–11.

27. Yoshimura S, Bondeson J, Brennan FM, Foxwell BM, Feldmann M. Role of NFkappaB in antigen presentation and development of regulatory T cells elucidated by treatment of dendritic cells with the proteasome inhibitor PSI. Eur J Immunol. 2001;31(6):1883–93.

28. Yoshimura S, Bondeson J, Foxwell BM, Brennan FM, Feldmann M. Effective antigen presentation by dendritic cells is NF-kappaB dependent: coordinate regulation of MHC, co-stimulatory molecules and cytokines. Int Immunol. 2001;13(5):675–83.

29. Gomez TS, Hamann MJ, McCarney S, Savoy DN, Lubking CM, Heldebrant MP, et al. Dynamin 2 regulates T cell activation by controlling actin polymerization at the immunological synapse. Nat Immunol. 2005;6(3):261–70.

30. Valitutti S. The Serial Engagement Model 17 Years After: From TCR Triggering to Immunotherapy. Front Immunol. 2012;3:272.

31. Ballabio A, Bonifacino JS. Lysosomes as dynamic regulators of cell and organismal homeostasis. Nat Rev Mol Cell Biol. 2020;21(2):101–18.

32. Jin J, Zhang H, Weyand CM, Goronzy JJ. Lysosomes in T Cell Immunity and Aging. Front Aging. 2021;2:809539.

33. Watts C. Lysosomes and lysosome-related organelles in immune responses. FEBS Open Bio. 2022;12(4):678–93.

34. Klein L, Munz C, Lunemann JD. Autophagy-mediated antigen processing in CD4(+) T cell tolerance and immunity. FEBS Lett. 2010;584(7):1405–10.

35. Chowdhury I, Siregar TAP, Haque MF, Ponpuak M. The final gate: bacterial subversion of autophagosome-lysosome fusion as a conserved immune evasion strategy. Crit Rev Microbiol. 2026;52(2):377–93.

36. Li Y, James SJ, Wyllie DH, Wynne C, Czibula A, Bukhari A, et al. TMEM203 is a binding partner and regulator of STING-mediated inflammatory signaling in macrophages. Proc Natl Acad Sci U S A. 2019;116(33):16479–88.

37. Lorincz P, Toth S, Benko P, Lakatos Z, Boda A, Glatz G, et al. Rab2 promotes autophagic and endocytic lysosomal degradation. J Cell Biol. 2017;216(7):1937–47.

38. Song W, Li D, Tao L, Luo Q, Chen L. Solute carrier transporters: the metabolic gatekeepers of immune cells. Acta Pharm Sin B. 2020;10(1):61–78.

39. Munz C. Antigen processing via autophagy--not only for MHC class II presentation anymore? Curr Opin Immunol. 2010;22(1):89–93.

40. Sanjabi S, Oh SA, Li MO. Regulation of the Immune Response by TGF-beta: From Conception to Autoimmunity and Infection. Cold Spring Harb Perspect Biol. 2017;9(6).

41. Ding M, Malhotra R, Ottosson T, Lundqvist M, Mebrahtu A, Brengdahl J, et al. Secretome screening reveals immunomodulating functions of IFNalpha-7, PAP and GDF-7 on regulatory T-cells. Sci Rep. 2021;11(1):16767.

42. Jenkinson G, Pujadas E, Goutsias J, Feinberg AP. Potential energy landscapes identify the information-theoretic nature of the epigenome. Nat Genet. 2017;49(5):719–29.

43. Xie H, Wang M, de Andrade A, Bonaldo Mde F, Galat V, Arndt K, et al. Genome-wide quantitative assessment of variation in DNA methylation patterns. Nucleic Acids Res. 2011;39(10):4099–108.

44. Litonjua AA, Lange NE, Carey VJ, Brown S, Laranjo N, Harshfield BJ, et al. The Vitamin D Antenatal Asthma Reduction Trial (VDAART): rationale, design, and methods of a randomized, controlled trial of vitamin D supplementation in pregnancy for the primary prevention of asthma and allergies in children. Contemp Clin Trials. 2014;38(1):37–50.

45. Mirzakhani H, Litonjua AA, McElrath TF, O’Connor G, Lee-Parritz A, Iverson R, et al. Early pregnancy vitamin D status and risk of preeclampsia. J Clin Invest. 2016;126(12):4702–15.

46. Aryee MJ, Jaffe AE, Corrada-Bravo H, Ladd-Acosta C, Feinberg AP, Hansen KD, et al. Minfi: a flexible and comprehensive Bioconductor package for the analysis of Infinium DNA methylation microarrays. Bioinformatics. 2014;30(10):1363–9.

47. Boyd SS, Hecker J, Mirzakhani H, DeMeo DL, Weiss ST, Halu A. Exploring associations between maternal vitamin D-binding protein, vitamin D, and offspring asthma/recurrent wheeze. Am J Clin Nutr. 2026;123(6):101188.

48. Teschendorff AE, Breeze CE, Zheng SC, Beck S. A comparison of reference-based algorithms for correcting cell-type heterogeneity in Epigenome-Wide Association Studies. BMC Bioinformatics. 2017;18(1):105.

49. Salas LA, Zhang Z, Koestler DC, Butler RA, Hansen HM, Molinaro AM, et al. Enhanced cell deconvolution of peripheral blood using DNA methylation for high-resolution immune profiling. Nat Commun. 2022;13(1):761.

50. Hoffman GE, Roussos P. Dream: powerful differential expression analysis for repeated measures designs. Bioinformatics. 2021;37(2):192–201.

51. Xu Z, Xie C, Taylor JA, Niu L. ipDMR: identification of differentially methylated regions with interval P-values. Bioinformatics. 2021;37(5):711–3.

52. Consortium EP, Moore JE, Purcaro MJ, Pratt HE, Epstein CB, Shoresh N, et al. Expanded encyclopaedias of DNA elements in the human and mouse genomes. Nature. 2020;583(7818):699–710.

53. Zou Z, Ohta T, Oki S. ChIP-Atlas 3.0: a data-mining suite to explore chromosome architecture together with large-scale regulome data. Nucleic Acids Res. 2024;52(W1):W45–W53.

54. Nurminen V, Neme A, Seuter S, Carlberg C. The impact of the vitamin D-modulated epigenome on VDR target gene regulation. Biochim Biophys Acta Gene Regul Mech. 2018;1861(8):697–705.

55. Seuter S, Neme A, Carlberg C. Epigenome-wide effects of vitamin D and their impact on the transcriptome of human monocytes involve CTCF. Nucleic Acids Res. 2016;44(9):4090–104.

56. Neme A, Seuter S, Carlberg C. Vitamin D-dependent chromatin association of CTCF in human monocytes. Biochim Biophys Acta. 2016;1859(11):1380–8.

57. Neme A, Seuter S, Carlberg C. Selective regulation of biological processes by vitamin D based on the spatio-temporal cistrome of its receptor. Biochim Biophys Acta Gene Regul Mech. 2017;1860(9):952–61.

58. Chen Y, Chen L, Lun ATL, Baldoni PL, Smyth GK. edgeR v4: powerful differential analysis of sequencing data with expanded functionality and improved support for small counts and larger datasets. Nucleic Acids Res. 2025;53(2).

59. Gene Ontology C. The Gene Ontology knowledgebase in 2026. Nucleic Acids Res. 2026;54(D1):D1779–D92.

60. Kanehisa M, Furumichi M, Sato Y, Matsuura Y, Ishiguro-Watanabe M. KEGG: biological systems database as a model of the real world. Nucleic Acids Res. 2025;53(D1):D672–D7.

61. Ragueneau E, Gong C, Sinquin P, Sevilla C, Beavers D, Grentner A, et al. The Reactome Knowledgebase 2026. Nucleic Acids Res. 2026;54(D1):D673–D81.

62. Wu T, Hu E, Xu S, Chen M, Guo P, Dai Z, et al. clusterProfiler 4.0: A universal enrichment tool for interpreting omics data. Innovation (Camb). 2021;2(3):100141.

63. Yu G, He QY. ReactomePA: an R/Bioconductor package for reactome pathway analysis and visualization. Mol Biosyst. 2016;12(2):477–9.

64. Traag VA, Waltman L, van Eck NJ. From Louvain to Leiden: guaranteeing well-connected communities. Sci Rep. 2019;9(1):5233.

65. Seale K, Horvath S, Teschendorff A, Eynon N, Voisin S. Making sense of the ageing methylome. Nat Rev Genet. 2022;23(10):585–605.

