## Additional file 2 for "Maternal vitamin D status during pregnancy is associated with reduced DNA methylation entropy and hypomethylation of regulatory regions related to immune tolerance"

### Slide 1
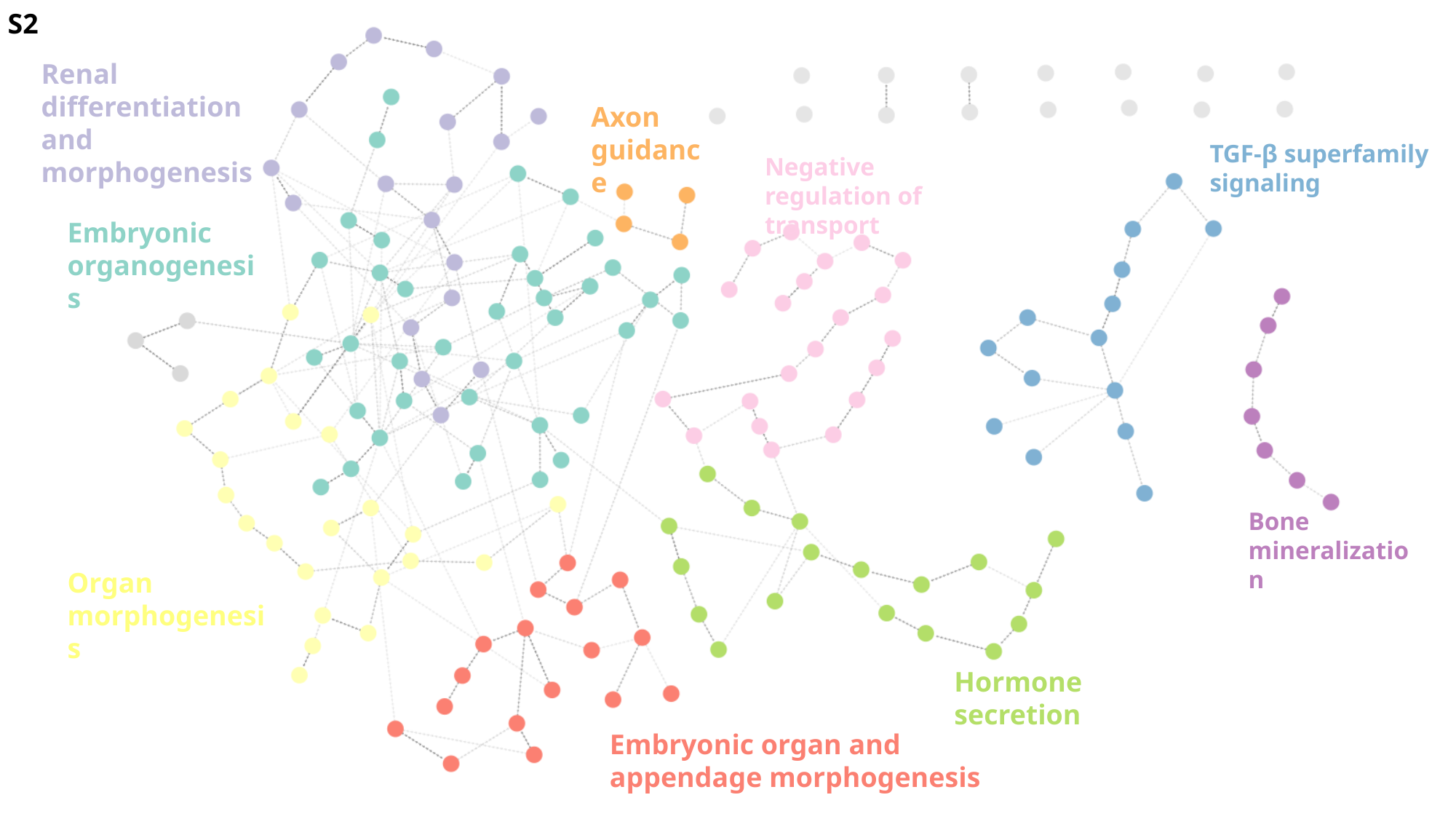

S2
Renal differentiation and morphogenesis
Axon guidance
TGF-β superfamily signaling
Negative regulation of transport
Embryonic organogenesis
Bone mineralization
Organ morphogenesis
Hormone secretion
Embryonic organ and appendage morphogenesis
