## Supplementary figures and images for "Maternal vitamin D status during pregnancy is associated with reduced DNA methylation entropy and hypomethylation of regulatory regions related to immune tolerance"

### Additional file 4

#### Slide 1
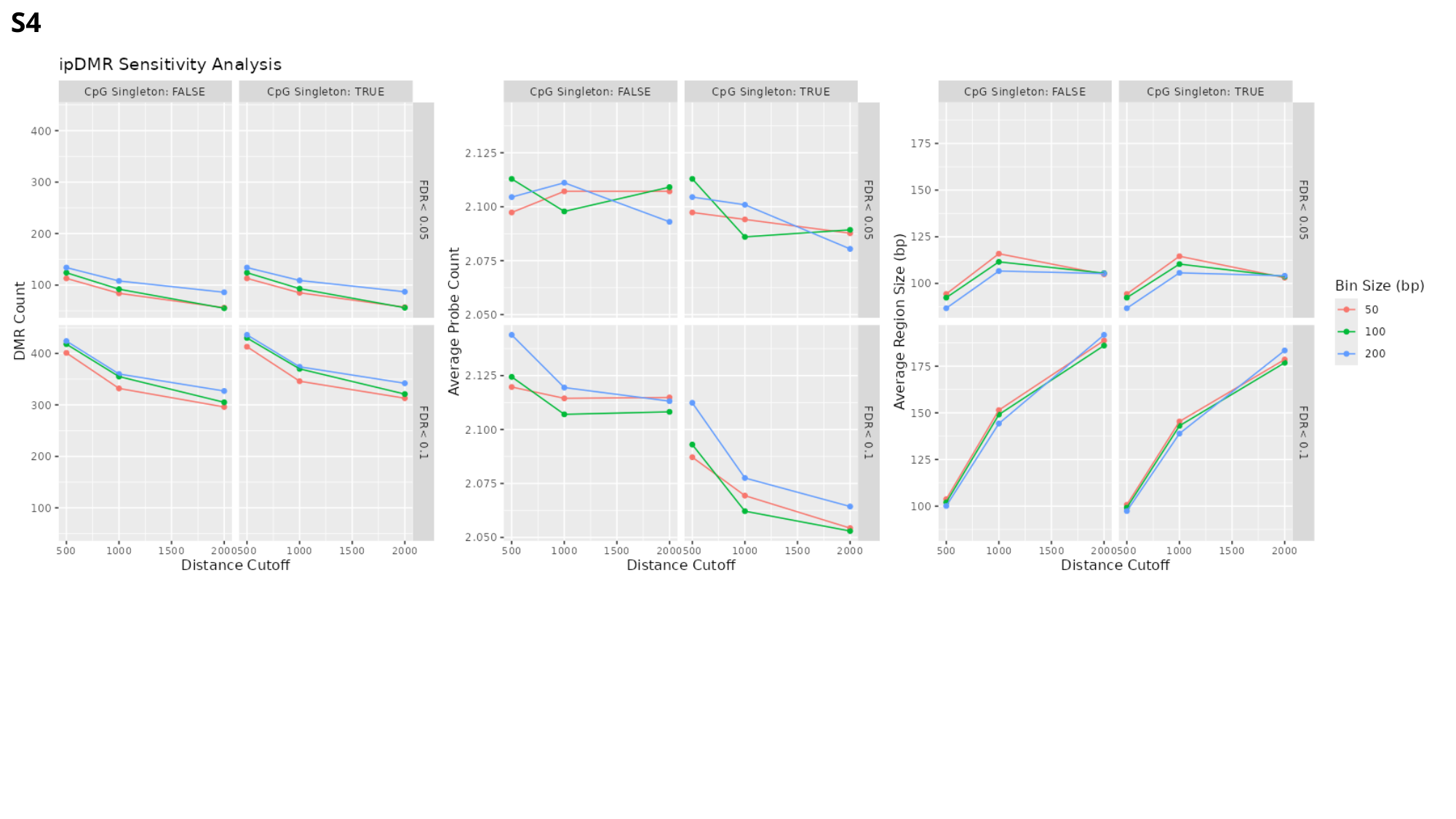

S4

### Additional file 5

#### Slide 1
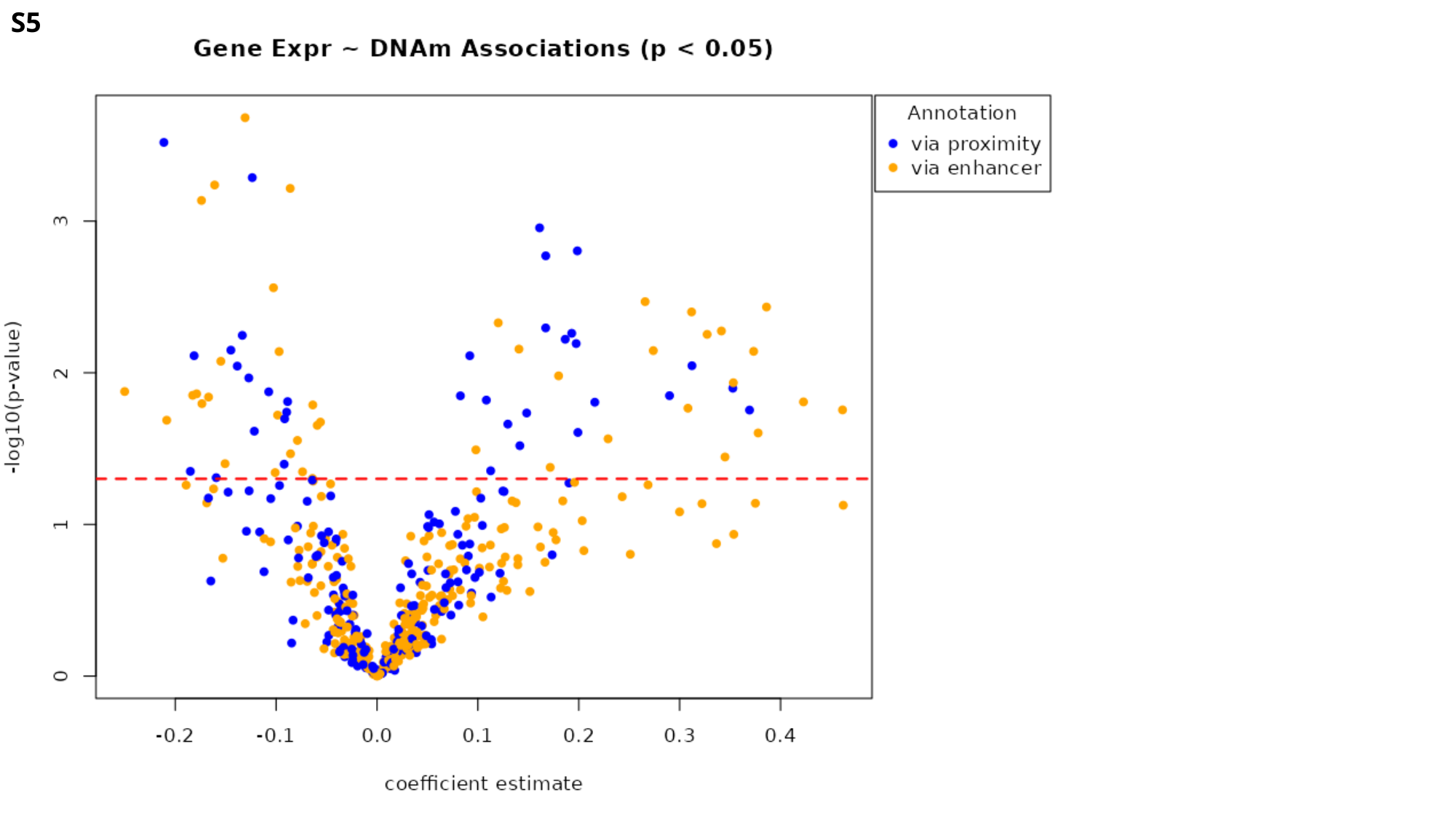

S5

### Additional file 6

#### Slide 1
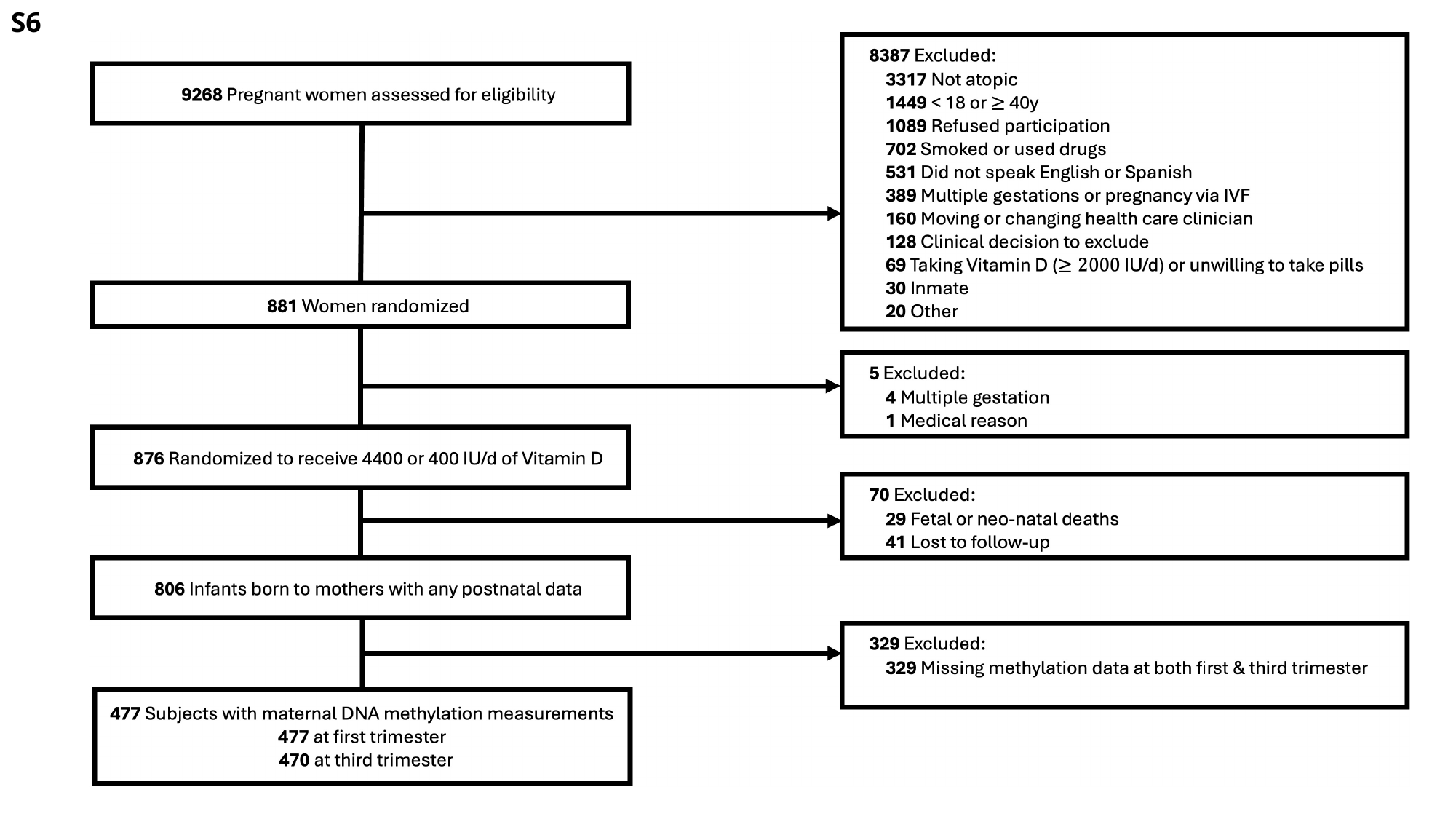

S6

### Additional file 7

#### Slide 1
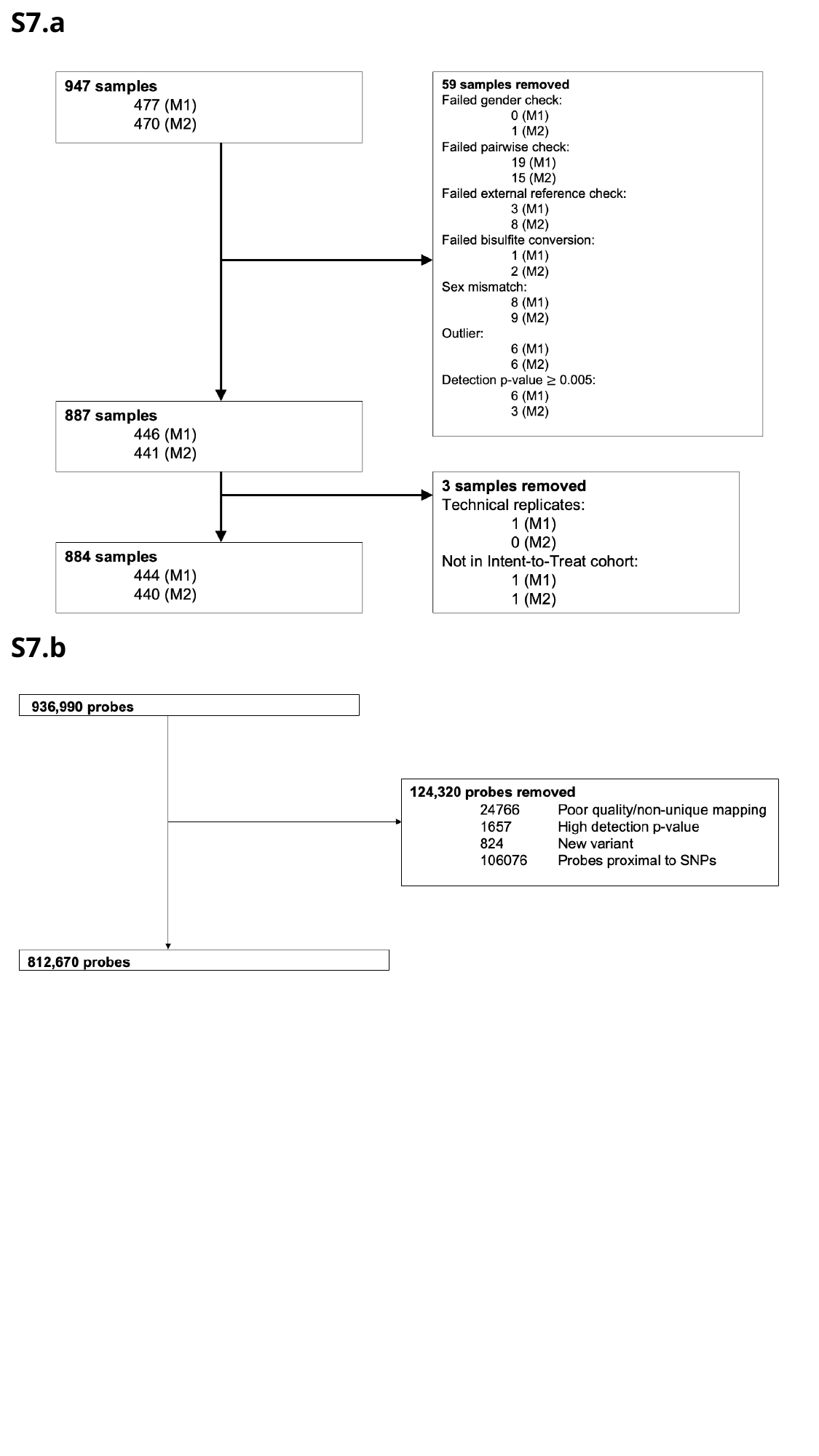

S7.a
S7.b
