## Additional file 1 for "Maternal vitamin D status during pregnancy is associated with reduced DNA methylation entropy and hypomethylation of regulatory regions related to immune tolerance"

Subset of VDAART Intent-to-Treat Cohort with DNAm Data

|  | **Time of Measurement** | |  | **p-value** |
| --- | --- | --- | --- | --- |
|  | **First trimester (N=444)** | **Third trimester (N=440)** | **Total (N=884)** |  |
| **Total 25OHD (ng/mL)** |  |  |  | <0.0001 |
| Mean (SD) | 24.2 (10.7) | 33.3 (14.5) | 28.7 (13.5) |  |
| Median [Min, Max] | 23.4 [5.85, 80.8] | 31.7 [6.08, 90.1] | 26.7 [5.85, 90.1] |  |
| Missing | 1 (0.2%) | 1 (0.2%) | 2 (0.2%) |  |
| **DBP (μg/mL)** |  |  |  | <0.0001 |
| Mean (SD) | 519 (130) | 673 (205) | 596 (188) |  |
| Median [Min, Max] | 516 [178, 1100] | 629 [295, 1750] | 569 [178, 1750] |  |
| Missing | 4 (0.9%) | 5 (1.1%) | 9 (1.0%) |  |
| **Gestational Age (weeks)** |  |  |  | <0.0001 |
| Mean (SD) | 14.1 (2.78) | 34.0 (1.60) | 24.0 (10.2) |  |
| Median [Min, Max] | 13.6 [2.14, 19.0] | 33.9 [29.1, 38.9] | 18.9 [2.14, 38.9] |  |
| Missing | 0 (0%) | 5 (1.1%) | 5 (0.6%) |  |
| **Season of Measurement** |  |  |  | 0.0005 |
| Fall | 87 (19.6%) | 126 (28.6%) | 213 (24.1%) |  |
| Spring | 142 (32.0%) | 95 (21.6%) | 237 (26.8%) |  |
| Summer | 110 (24.8%) | 124 (28.2%) | 234 (26.5%) |  |
| Winter | 105 (23.6%) | 95 (21.6%) | 200 (22.6%) |  |
